# Sexual orientation inequalities in mental health across adolescence and early adulthood: exploring the contribution of cumulative experiences of bullying and victimisation over time

**DOI:** 10.64898/2026.07.31.26359405

**Authors:** Amal R. Khanolkar, Laia Becares

## Abstract

**Background:** Sexual minority ([SM] or LGB+) adolescents experience mental health (MH) inequities and disproportionately high rates of bullying and victimisation. Less is known about the cumulative effects of bullying and victimisation across adolescence on MH in early adulthood, or the moderating role of loneliness and social support in this relationship.

**Objective:** To investigate cumulative effects of bullying and victimisation in adolescence on MH in early adulthood, including the role of loneliness/social support.

**Methods:** Drawing on a UK-wide nationally representative sample of individuals (N=12,872/LGB+ 22%) with self-reported bullying and victimisation assessed at ages 11/14/17/23, and MH (psychological distress/self-harm/attempted suicide) at age 23. Logistic models assessed separate associations between cumulative bullying or victimisation (0, 1, ≥2times) and MH, and differences by sexual identity (using appropriate interactions), with adjustment for loneliness and social support.

**Findings:** Across adolescence, SM individuals experienced higher prevalence of cumulative victimisation (3 times; gay/lesbian:22%, bisexual:22% vs heterosexual:9%) and bullying (2 times; gay/lesbian:35% and bisexual:27% vs heterosexual:18%).

Models revealed substantial inequalities with higher proportions of SM individuals reporting self-harm compared to heterosexual peers with the same levels of experienced victimisation. Among those reporting victimisation 1 and ≥2 times, 15% (95% CI 13-16%) and 23% (21-25%) of heterosexual individuals reported self-harm, increasing to 41% (33-50%) and 53% (47-59%) for bisexual, and 24% (10-39%) and 49% (40-59%) for gay/lesbian individuals, and 24% (11-38%) and 53% (39-67%) for other SM identities, respectively.

Similarly, for heterosexual individuals experiencing victimisation 1 and ≥2 times, 6% (5-8%) and 13% (11-14%) reported attempted suicide respectively. This increased to 21% (13-29%) and 38% (33-44%) for bisexual, and 17% (6-29%) and 34% (24-44%) for gay/lesbian individuals experiencing victimisation 1 and ≥2 times, respectively.

A key finding was that adjustment for loneliness and lack of peer/family support reduced the proportions reporting adverse mental health by 15-45% among SM but not heterosexual individuals. Similar patterns by sexual identity were found for bullying and all indicators of MH.

**Conclusions:** SM individuals experience higher levels of cumulative bullying and victimisation across adolescence, which is associated with substantially higher rates of mental ill-health in early adulthood. Experiences of loneliness explain MH inequalities to greater extent in SM youth compared to heterosexual peers.

**What is already known on this topic:** Evidence shows that sexual minority adolescents report both higher levels of bullying and worse mental health compared to heterosexual peers. However, no study has comprehensively examined the cumulative impact of types of bullying (e.g., bullied by peers/siblings or victimised by adverse actions) across adolescence and mental health inequalities by sexuality in early adulthood when individuals navigate access to the labour market and higher education. The role of modifiable factors like social isolation and support remains less understood.

**What this study adds:** The compounding double disadvantage of sexual minority identity intersectionally layered with cumulative bullying histories across adolescence drive substantial inequities in mental health in early adulthood. Further, these inequities were substantially explained by loneliness and poor social support in sexual minority but not heterosexual young adults. This is the first study to demonstrate the long-term impact of different types and cumulative experiences of bullying in sexual minority sub-groups in a nationally representative sample.

**How this study might affect research, practice or policy:** Reducing social isolation and improving access to peer support can substantially reduce mental health problems including attempted suicide in sexual minority young adults.

## Introduction

People who self-identify as lesbian, gay, bisexual, queer or any other non-heterosexual identity ([LGB+], hereon sexual minority people) experience persistent and substantially higher risk for several mental health problems including depression, self-harm and suicidality across the lifecourse^1^. These inequities in mental health are attributed to experiences of stress, violence, stigma and discrimination associated with sexual minoritized identities^2–4^. Chronic exposure to these minoritised related discrimination and stressors creates a stressful environment leading to worse mental health as exemplified in Minority stress theory^5^. Sexual minority individuals experience minoritised identity-associated discrimination and stigma in addition to other forms of discrimination experienced by heterosexual peers^4^. These minoritised associated discrimination also lead to higher levels of adverse health behaviours which are both coping mechanisms to live with adverse discrimination and independently associated with poor mental health^2 6^. Experiences of discrimination occur over the life course of sexual minority people, and during the early life course and adolescence often take the form of bullying and victimisation.

There is no legal definition of what constitutes bullying in the UK, but it is commonly defined as intentional and repeated behaviour that emotionally and/or physically hurts someone and includes in-person (traditional bullying) and online (cyberbullying)^7^. Bullying is common in childhood especially in early to mid-adolescence^8 9^. The Crime Survey for England and Wales (CSEW) 2023 and the National Behaviour Survey (NBS) 2024-25 are critical sources on the prevalence and types of bullying experienced by UK children^10 11^. Most recent survey data suggest that 20-35% of children aged 7-15 years experienced bullying in the past year with prevalence similar in both sexes and in-person being more common than online bullying. When estimates from the nationally representative CSEW are extrapolated nationally, this corresponds to 1.5 million and 847,000 children who reported in-person and online bullying, respectively^11^. Appearance, having a disability, and protected characteristics like sexual identity, ethnicity, faith and nationality are the most common reasons for bullying in adolescence^11^. In some studies, ‘bullying’ and ‘victimisation’ are distinguished to capture the specific source and nature of peer aggression. While bullying often denotes the general experience of being picked on by peers or siblings, victimisation typically refers to a broader range of specific acts, such as physical assault or relational aggression, allowing for a more granular analysis of cumulative adolescent adversity. Nonetheless, these variables often overlap to create a compounding effect on mental health, and specifically in sexual minority adolescents^9^. Bullying and victimisation are established causal risk factors for poor mental health including over two times higher risk for depression and anxiety, non-suicidal self-harm and attempted suicide and over six times higher risk for PTSD^12^. Bullying and victimisation are both associated with poorer educational outcomes, crime and substance use, behavioural issues and is recognised as a public health concern^9 13 14^.

Substantial evidence shows that sexual minority young people are more than twice as likely to experience both in-person and online bullying and victimisation compared to heterosexual peers^9 15–17^, with some evidence that bisexual youth face higher levels of bullying, especially online bullying compared to gay/lesbian peers^18^. Further, sexual minority youth are more likely to experience frequent and severe bullying compared to heterosexual peers which partially contributes to widening sexual identity inequalities in mental and sexual health, life satisfaction and wellbeing across adolescence^19^. Sexual minority youth are at increased risk for all types of bullying and victimisation, including physical, sexual, verbal, online and homophobic bullying and social isolation. Nearly 50% of sexual minority youth report facing bullying and discrimination (or homophobic bullying) associated with their sexual minority identity in schools and universities in the UK leading to feeling unsafe^20^. More concerning is that ∼50% never report being bullied and a staggering 72% said that staff failed to act or responded badly^20^. The rates of homophobic (or LGBT-phobic) bullying and victimisation experiences vary substantially by study sample, size, representativeness and country, but are substantially high in all studies conducted to date. Further, sexual minority adolescents are more likely to experience social isolation and reduced peer support both as part of homophobic bullying and as a consequence potentially exacerbating poor mental health^21 22^. A key point from the existing literature is that the detrimental effects of bullying and victimisation on mental health while observed in both heterosexual and sexual minority individuals, is much worse in the latter^15^. This is concerning as the detrimental effects of bullying in adolescence persist into adulthood exacerbating other life challenges.

Despite substantial evidence on sexual identity differences in bullying and victimisation and emerging evidence in how these inequalities impact mental health, these studies are limited by cross-sectional design, smaller and/or non-probability samples, combining sexual minority groups together and/or examining associations with single mental health problems. In the UK, longitudinal studies on sexual identity differences on the impact of bullying on mental health are limited and restricted to adolescence only. Findings indicate that sexual minority adolescents are more likely to experience long-lasting victimisation associated with worse mental health in late adolescence, and experienced greater increases in depressive symptoms across adolescence which was partially explained by higher levels of bullying in early adolescence^23 24^. A critical gap in knowledge is sexual identity differences in the types of and cumulative bullying and victimisation experiences across adolescence and subsequent impact on mental health in early adulthood – a critical time when individuals navigate access to higher education and the labour market. This is imperative to study as the impact of cumulative experiences of bullying and victimisation on mental health is established across the lifecourse and in marginalised groups like ethnic minority groups^25 26^.

Further, the role of mediating and moderating factors, including experiences of social isolation and peer support in the impact of bullying on mental health is poorly understood in sexual minority individuals, and especially in early adulthood. Sexual minority individuals report higher levels of loneliness or social isolation across the lifecourse, which mediates subsequent depression to a greater extent in sexual minority compared to heterosexual individuals^27^. Understanding sexual identity differences in the long-term effects of cumulative bullying and victimisation on mental health including the role of loneliness and social support will aid in designing policies that can reduce persisting mental health inequities.

This study contributes to wider understanding of the drivers of mental health inequalities experienced by sexual minority people by examining the associations between cumulative experiences of bullying and victimisation across adolescence and mental health problems in early adulthood (age 23), and whether these associations differed by sexual identity in a large nationally representative UK-wide sample. It also examines the contribution of experiences of loneliness and lack of social support in explaining associations between bullying/victimisation and mental health outcomes in sexual minority individuals, compared to heterosexual peers.

## Methods

### Study design and participants

The Millennium Cohort Study (MCS) is a UK birth cohort study following 19,519 children born at the start of the millennium (2000–2002). Participants have been followed over eight sweeps to date (9 months, 3, 5, 7, 11, 14, 17 and 23 years). Detailed information on the study, sampling and survey design can be found at: https://cls.ucl.ac.uk/cls-studies/millennium-cohort-study/. Briefly, a nationally representative birth cohort from across the UK was recruited to the MCS and included children living in non-household situations and those not born in the UK but lived in the country at recruitment. The sampling strategy was to recruit 100% of eligible children living in geographically defined areas of residence (the boundaries of electoral wards as defined before the 2001 national census) during the eligible period. A stratified cluster sampling design was used to ensure adequate representation of families living in disadvantaged areas and from ethnic minority groups and the overall response rate was 72%. Children living in disadvantaged areas, those of ethnic minority backgrounds and growing up in the smaller nations of the UK were intentionally over-sampled. Attrition across sweeps was predicted by single-parent families, lower-income occupation and lower educational level, Black ethnicity and male sex. The STrengthening the Reporting of OBservational studies in Epidemiology (STROBE) were followed in the reporting of this manuscript.

Data for this study was primarily from the ages 11 (MCS 5), 14 (MCS 6), 17 (MCS 7) and 23 (MCS 8) sweeps, which collected data on self-reported bullying and victimisation, and coincides with sexual identity development and exploration.

### Sexual identity

The main exposure of interest was sexual identity which was self-reported and assessed at age 17. Participants could choose from one of eight options (listed in Supplemental Table 1) and were categorised into the following: (1) Completely heterosexual, (2) Mainly heterosexual, (3) Bisexual, (4) Gay or lesbian and (5) Other sexual minority identities. The other sexual minority identities category included those who chose the options ‘other’, ‘do not know’, and ‘prefer not to say’.

### Cumulative experiences of frequent bullying and victimisation across adolescence

At ages 11 and 14, bullying was assessed by two identical questions *“How often do your brothers or sisters hurt you or pick on you on purpose?”* and *“How often do other children hurt you or pick on you on purpose?”* A third question *“How often do other children bully you online”* was additionally asked at age 14 only. All three questions had the same seven options: most days, about once a week, about once a month, every few months, less often and never. For each question, we created a binary variable that grouped adolescents into 1. Frequent bullying (most days and about once a week) or 2. Less frequent/never (all other options). Next, we created an overall bullying variable at each age to indicate those who reported bullying vs those who did not. Finally, we created an overall cumulative bullying variable which indicated the number of experiences of bullying at different timepoints in adolescence with the categories 1.Never bullied, 2.Bullied once (either age 11 or 14) and 3.Bullied twice (both timepoints or ages 11 *and* 14).

Victimisation was assessed at ages 14, 17 and 23. At each sweep participants were asked *“In the past 12 months has anyone done any of these things to you?”* and participants could choose one or more options from 1. Insulted/threatened/shouted at, 2. Experiences of physical violence, 3. Hit or used a weapon, 4. Had something stolen and 5. Been sexually assaulted. At each age we created a binary variable to indicate no experiences of victimisation vs any (≥1) experiences of victimisation. We created an overall cumulative victimisation variable which indicated the number of experiences of victimisation at different timepoints in adolescence and early adulthood with the categories 1.Never victimised, 2.Victimised once (either age 14, 17 or 23) and 3.Victimised ≥2 times (i.e., at any two or more ages or timepoints).

This cumulative measure of bullying and victimisation is based on a measure of cumulative exposure to racial discrimination used in previous studies, including with the MCS^15 26^.

### Mental health outcomes at age 23

We included three indicators of mental health: psychological distress, self-harm, and attempted suicide. Psychological distress (symptoms of depression/anxiety) was assessed via the validated 6-item Kessler Psychological Distress Scale (K6)^28^. The K6 is a self-reported questionnaire comprising six questions or items (for e.g., *“How often have you felt hopeless?”*) with each item rated on a 5-point Likert scale, with total scores ranging from 0 to 24 (higher scores indicating higher probability of mental health problems). The K6 captures symptoms in the preceding 30-days and is routinely used in population-based surveys.

Self-harm in the previous year and lifetime attempted suicide were self-reported. Self-harm was captured with a series of questions that asked participants about self-harming actions (like cutting/stabbing, burning, bruising/pinching, taking an overdose of tablets, pulling out hair or hurt one-self in any other way). We created binary variable indicating no self-harm vs any kind of self-harm. Attempted suicide was captured by asking participants: *‘’Have you ever hurt yourself on purpose to end your life?’’* Responses were categorised as no vs yes.

### Loneliness and social support

The validated 3-item short version of the UCLA Loneliness Scale assessed loneliness^29^. This scale comprises three questions on lack of companionship, feeling left out and feeling isolated which assesses three specific dimensions of loneliness: relational connectedness, social connectedness, and perceived isolation. Each question captures responses on a 3-point Likert scale with total scores ranging from 3 to 9. Higher scores indicate higher levels of loneliness. We used the validated cut-off of 6 to group participants into “not lonely” (3-5) and “lonely” (6-9).

To measure social support, we used questions that asked participants if they had individuals in their current relationships (including friends and family) who made them feel safe, secure and happy. We created a binary variable which indicated those who answered 1. Very true and partly true vs 2. Not true at all.

### Covariates

Covariates included ethnicity, assigned sex at birth, and parental income. Ethnicity was self-reported by participants at age 14. Any missing information on ethnicity was first replaced with self-reported data from age 11 and subsequently by parent reported ethnicity from the age 3 sweep. Assigned sex at birth was assessed at the birth sweep. For analyses, ethnicity was coded as a binary variable (White vs non-White). Parental income was used as an indicator of socioeconomic position ascertained at age 11. Household income (Organisation for Economic Co-operation and Development UK) was categorised into equalised quintiles (where quintiles 1 and 5 represent the lowest and highest income quintiles respectively). Missing data on parental income was first replaced by data from the age 14 followed by the age 3 sweeps.

Complete details, the original questions, all component items of scales, questions across all sweeps and how these were operationalized for analyses (including references) are listed in Supplementary Table 2.

### Missing data

As with most longitudinal cohort studies, the MCS has experienced loss-to-follow-up. Missing data in the four sweeps (ages 11 to 23) was addressed using multiple imputation with chained equations assuming data missing at random ([MAR] and including 40 imputations)[44]. The MAR mechanism (often largely untestable) implies that systematic differences between the missing values and the observed values can be explained by observed data, a valid assumption in the British birth cohorts given the rich data available from birth. In addition to ethnicity and socioeconomic indicators, it is reasonable to assume that mental health indicators, bullying and victimisation are also MAR given their significant association with many variables including the wide range of health outcomes and health-risk behaviours available in addition to being collected across multiple sweeps[20]. The main purpose of imputation was to address missing data in mental health indicators, bullying and victimisation. If a study participant had data on at least one mental health indicator (psychological distress, self-harm, attempted suicide recorded at any age between 11 and 23) *and* bullying/victimisation (recorded at any age between 11 and 23), then the participant was included in the sample, and any missing mental health and bullying/victimisation data was imputed. Ethnicity and parental income were available >98% of study participants and all participants had data on sex at birth. For most variables, missing data was <22% and with higher levels of missingness observed for alcohol consumption (44%), mental wellbeing (32%), BMI (37%), loneliness (32%) and mental health outcomes (∼30%) at age 23. We included a rich set of auxiliary variables including BMI, self-reported long-term illness, psychological distress, self-harm, attempted suicide, mental wellbeing, self-esteem, social support, number of close friends and life satisfaction (wherever available), smoking and alcohol consumption at ages 17 and 23, parental educational level at age 14, parental report of psychosocial distress in children at ages 11 and 14, whether been to university and sought mental health support at age 23 to help strengthen the quality of imputed data. Auxiliary variables with stronger associations with incompletely observed variables or the probability of data being missing, increases the potential for reducing bias^30^. Further, including both baseline (age 11) and longitudinal auxiliary variables (BMI, mental health, life satisfaction, long-term illness, mental wellbeing) increases the efficiency of imputing missing data^30^. Validity of the imputed dataset was verified by comparing all variables (means and SDs for continuous variables and proportions for categorical variables) between the complete case (CC) and imputed data (Supplemental Table 2). Post-imputation, the final study sample included 12,782 individuals. Proportions of individuals across the categories of key variables were very similar between CC and imputed samples (Supplemental Table 2).

#### Statistical analyses

All statistical analyses were conducted on the imputed dataset using Stata V19 (College Station, TX, USA).

Initial descriptive analysis included estimating the prevalence and distribution by sexual identity of bullying and/or victimisation, mental health outcomes, and covariates.

We first examined associations between sexual identity and the cumulative indicators of bullying (0, 1 or 2 times [at both ages 11 and 14]) and victimisation (0, 1 and ≥2 times [i.e., at all ages 14, 17 and 23]) using multinomial logistic regression models. Models were run unadjusted, and adjusted for assigned sex at birth, ethnicity and parental income.

Next, we separately examined associations between sexual identity and each of the mental health outcomes (psychological distress, self-harm and attempted suicide). As we are interested in whether higher levels of bullying and victimisation are associated with worse mental health outcomes in sexual minority young adults compared to heterosexual peers, these models included interaction terms between sexual identity and bullying or victimisation. We ran four models with adjustment as follows: Model 1 (unadjusted); Model 2 adjusted for assigned sex at birth, ethnicity and parental income; Model 3 additionally adjusted for loneliness; and Model 4 additionally adjusted for social support. To aid understanding of the interaction terms and inequalities in mental health outcomes, we estimated predicted probabilities (or *margins*) for each outcome across each of the fifteen categories (generated by the interaction terms between the bullying or victimisation and five sexual identity categories) using the *‘margins’* command in Stata. We estimated predicted probabilities for models 1 and 4 to show the change in probabilities after adjusting for covariates and how these differed across sexual identity groups. Predicted probabilities were visualised to better understand the interactions terms between bullying, victimisation and sexual identity variables. To account for the stratified cluster design of the MCS and attrition over time, all regression analyses were weighted with non-response weights from the birth sweep (using Stata’s *‘svy’* command for survey data).

#### Sensitivity analysis

We also ran regression models for the associations between sexual identity and cumulative bullying or victimisation, and sexual identity and each mental health outcome including interaction terms between sexual identity and bullying/victimisation variables (fully adjusted model only) in the CC sample. This was done to examine whether associations were similar between the imputed and CC samples.

## Results

All sexual minority groups had significantly higher prevalence of mental health problems compared to heterosexual peers (Table 1). For example, psychological distress and self-harm were more than twice as high in bisexual (39.5%) and gay/lesbian (38.8%) vs heterosexual (16.9%) groups. Greater proportions of sexual minority groups reported cumulative bullying and cumulative victimisation (e.g., mainly heterosexual: 51.5%, bisexual: 59.2%, gay/lesbian: 56.4% vs 35.5% in heterosexual groups). All sexual minority groups were significantly more likely to report feelings of loneliness (49.9% in bisexual, 48.9% in gay/lesbian, and 61.5% in other sexual minority groups vs. 31.9% in heterosexual individuals) and lack of social support (Table 1).

**Table 1.**
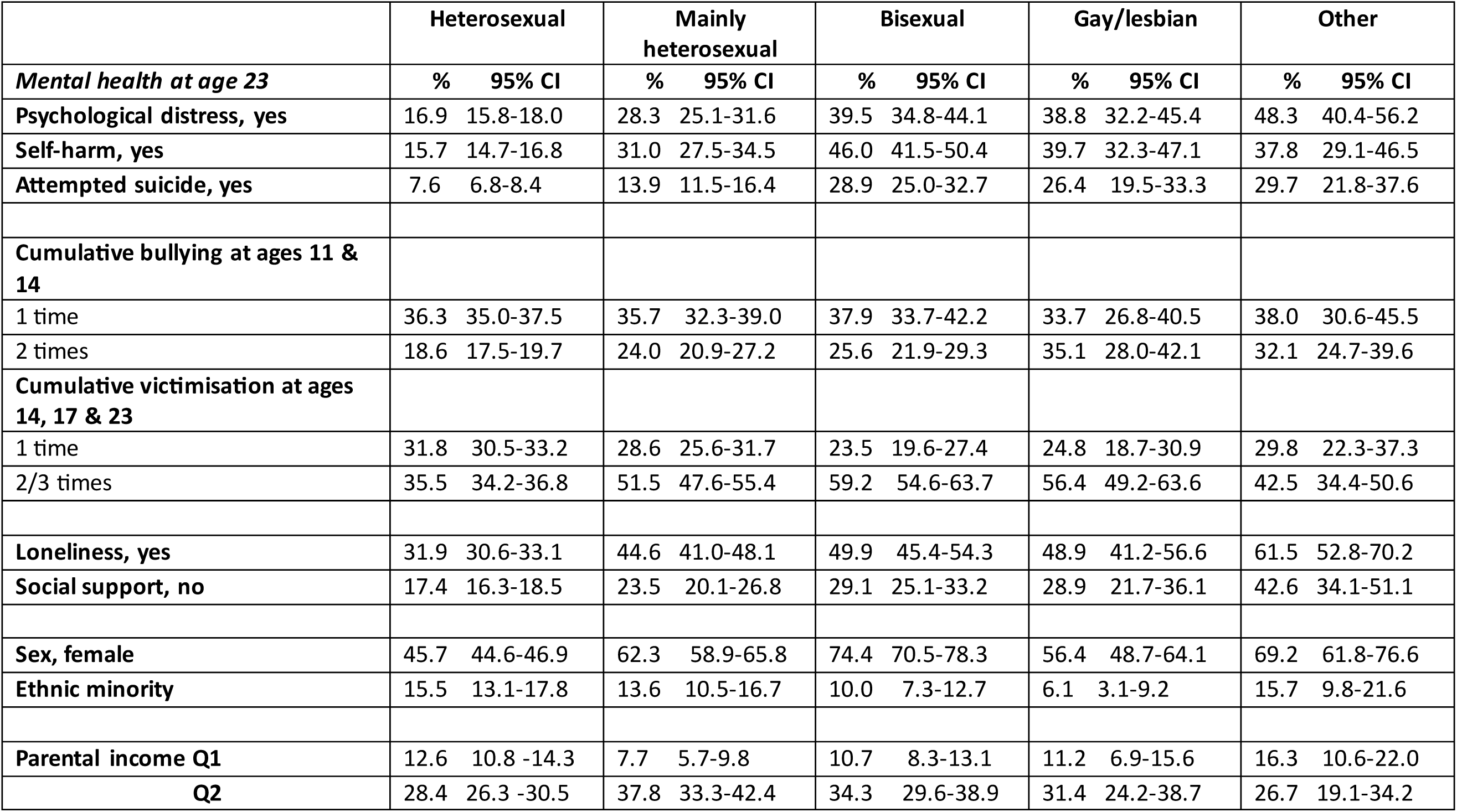
Descriptive statistics of variables of interest in 12,782 participants from the Millennium Cohort Study. All numbers are percentages (% [95%])

### Sexual identity inequalities in cumulative bullying and victimisation

Supplemental Table 3 and Figure 1 display estimates from multinomial logistic regression models on associations between sexual identity and experiencing cumulative bullying and victimisation across adolescence. Compared to heterosexual individuals, all sexual minority groups were significantly more likely to experience cumulative bullying (i.e., bullied at both ages 11 and 14) and cumulative victimisation (i.e., victimised at two or more age points between ages 14-23). For example, bisexual (adjusted RRR 3.18, 95% CI 2.44-4.15), gay/lesbian (aRRR 2.68, 1.79-4.00) and mainly heterosexual (aRRR 2.43, 1.95-3.02) individuals were 2.5-3 times were more likely to experience cumulative victimisation over time, compared to heterosexual peers. Similar pattern of associations was found for bullying, with effect estimates being stronger for the other sexual identities group compared to those observed for victimisation.

**Figure 1.**
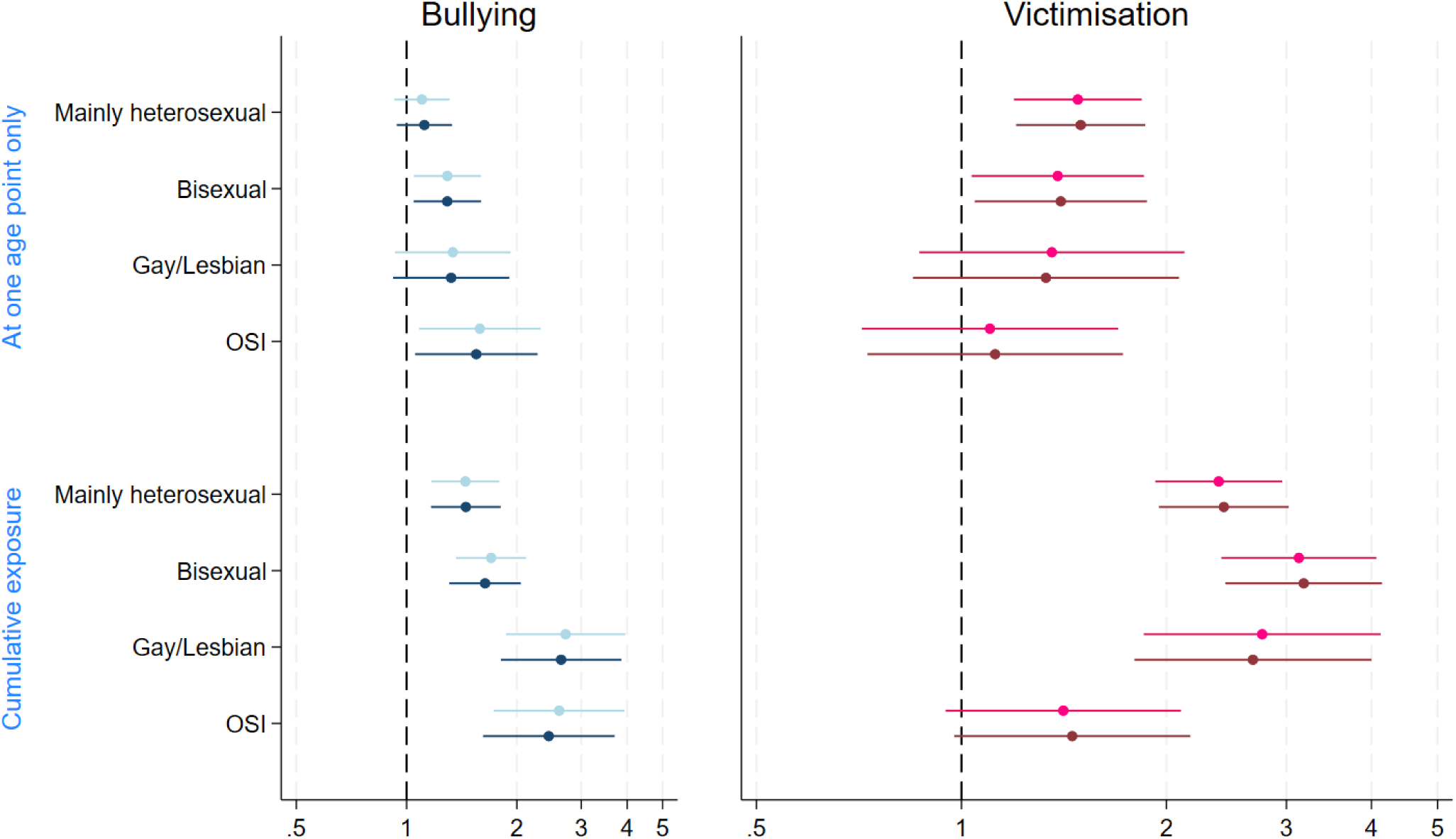
Associations between sexual identity and cumulative bullying or victimisation across adolescence and early adulthood in 12,872 individuals from the Millennium Cohort Study.

### Cumulative bullying and mental health

In the full sample, cumulative bullying (2 times across 2 time periods) and cumulative victimisation (2 or 3 times across 2 or 3 time periods) were associated with higher odds of poor mental health across the three outcomes (e.g., unadjusted OR 1.89, 1.57-2.28 for bullying and psychological distress, OR 1.92, 1.49-2.48 for bullying and attempted suicide) compared to never being bullied or victimised (Supplemental Tables 4 and 5). Adjustment for loneliness (Model 3) substantially attenuated estimates for all three mental health outcomes in models for both bullying and victimisation. Further adjustment for social support also attenuated estimates for all three outcomes but to a lesser degree. Cumulative victimisation was associated with larger odds ratios for all three outcomes compared to cumulative bullying (e.g., for attempted suicide, victimisation; adjusted OR 3.52, 2.57,4.82 vs bullying; aOR 1.57, 1.20,2.07).

Supplemental Tables 6 and 7 (Figures 2 and 3) display the crude and adjusted predicted probabilities for mental health outcomes in relation to intersectional sexual identities and cumulative bullying and victimisation, respectively. Compared to those never bullied, individuals who experienced bullying reported increased psychological distress. Further, there was a clear gradient, with proportions of individuals reporting psychological distress substantially increasing with greater number of bullying experiences at both ages 11 and 14, a pattern observed in all sexual minority groups. For example, among those who did not experience bullying, 14% of heterosexual, 22% of mainly heterosexual, 36% of bisexual, 31% of gay/lesbian and 39% of other sexual identity individuals reported psychological distress. These proportions increased substantially to 23% of heterosexuals, 37% for mainly heterosexual, 45% of bisexual, 47% of gay/lesbian and 57% of other sexual identity groups among those who experienced cumulative bullying. Similar patterns were observed for self-harm and attempted suicide (e.g., 16% of heterosexual who experienced bullying at one timepoint reported self-harm with corresponding estimates being 33% in mainly heterosexual, 47% in bisexual, 34% in gay/lesbian, and 35% in other sexual identity groups. These proportions increased to 20% in heterosexual, 38% in mainly heterosexual, 48% in gay/lesbian and 44% in other sexual identity groups who experienced cumulative bullying, i.e., at both ages 11 and 14).

**Figure 2.**
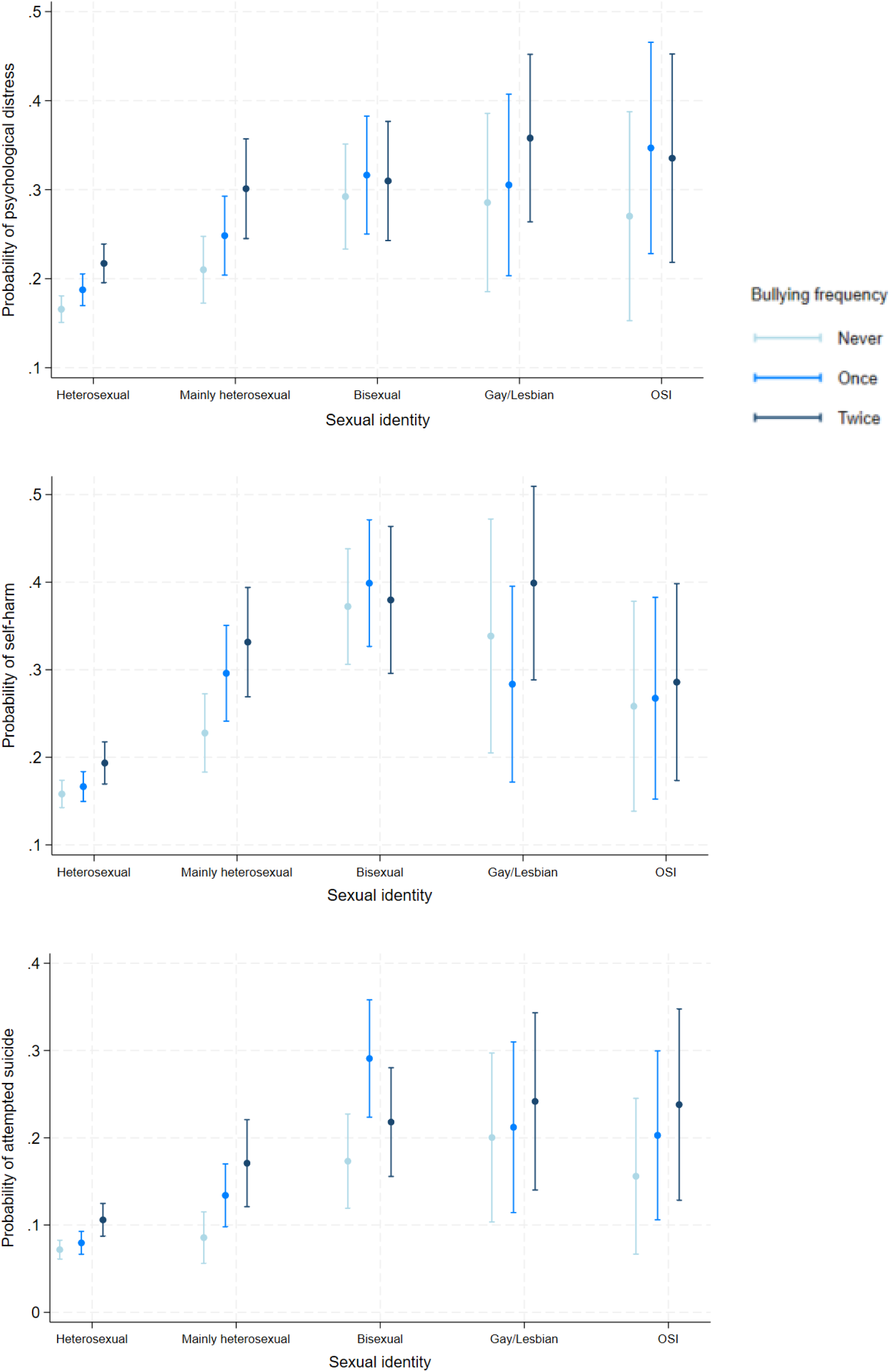
Predicted probabilities for mental health at age 23 based on bullying in adolescence in 12,782 individuals from the Millennium Cohort Study.

**Figure 3.**
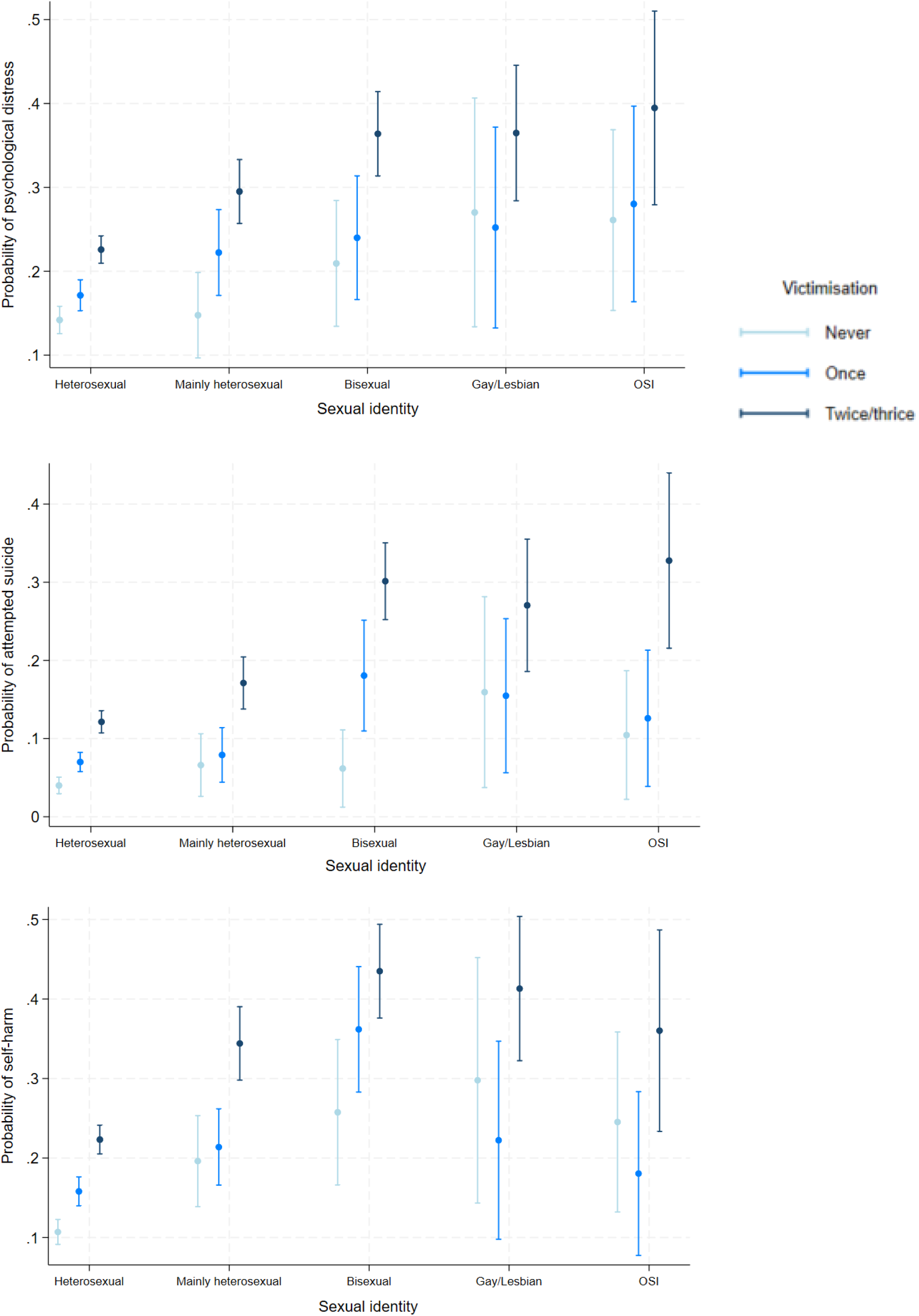
Predicted probabilities for mental health at age 23 based on victimisation in adolescence in 12,782 individuals from the Millennium Cohort Study.

### Cumulative victimisation and mental health

Across all sexual identity groups, experiences of victimisation were consistently associated with higher probabilities of reporting poor mental health across the three outcomes. Similar to bullying, sexual minority individuals reported higher baseline levels of adverse mental health and had higher proportions of individuals reporting adverse mental health compared to heterosexual individuals with the same frequency of victimisation. However, sexual minority individuals experienced a steeper gradient of adverse mental health as cumulation of victimisation increased. For example, proportions of individuals reporting psychological distress among those who did not experience victimisation were heterosexual: 11%, mainly heterosexual: 14%, bisexual: 25%, gay/lesbian: 26% and other sexual identities: 35%, which increased to heterosexual:15% and 24%, bisexual: 29% and 48%, gay/lesbian:29% and 47% and other sexual identities: 41% and 62%, among those who experienced victimisation 1 and ≥2 times (cumulative exposure to victimisation at 2 or 3 times between ages 14 and 23), respectively.

For both bullying and victimisation, individuals identifying as mainly heterosexual reported adverse mental health outcomes at rates that fell consistently between those for heterosexual and bisexual/gay/lesbian/ other sexual identity groups, i.e. with a gradient. Furthermore, the other sexual identity group reported the highest proportions of individuals with psychological distress (e.g., unadjusted proportions of 62%, 50-74%) and attempted suicide (48%, 35-61%) at the highest frequencies of bullying and victimisation. However, confidence intervals did overlap with those of bisexual and gay/lesbian individuals.

### The role of loneliness and social support

A key finding was that adjustment for loneliness and social support substantially reduced the predicted probabilities or proportions of individuals reporting adverse mental health in all sexual minority groups compared to heterosexual individuals. In fact, adjustment for loneliness and social support had little to no impact on predicted probabilities for the heterosexual group. For example, for self-harm, change in predicted probabilities for cumulative bullying were as follows: heterosexual, 20% (17-22%) to 19% (17-22%), mainly heterosexual, 38% (32-45%) to 33% (27-39%), bisexual, 49% (39-58%) to 38% (30-46), gay/lesbian, 48% (36-59%) to 40% (29-51%), other sexual identities, 44% (30-59) to 29% (17-40), for model 1 to model 4, respectively. A similar pattern was observed for predicted probabilities for psychological distress and attempted suicide.

Notably, adjustment for loneliness and support made little to no change in the proportions of heterosexual individuals reporting adverse mental health but substantially attenuated the proportions of sexual minority individuals reporting adverse mental health, especially among the bisexual, gay/lesbian and other sexual identity groups.

The observed nonsignificant interaction terms (p>0.05, Supplemental Tables 4 and 5) indicate that cumulative bullying/victimisation and sexual identity function as distinct, parallel risks for mental health. However, the predicted probabilities reveal a stark additive compounding of absolute risk, with chronic victimisation driving adverse mental health to their highest levels across all sexual minority groups.

### Sensitivity analyses

Patterns of associations including strength of effects sizes were similar in models restricted to CC samples (Supplemental Tables 7 and 8). Few effect estimates were not statistically significant, but the strength and direction of associations were consistent.

## Discussion

This study is the first to provide evidence on the longitudinal association between cumulative exposure to bullying and victimisation over time, and their association with worsening of mental health in sexual minority adolescents. Using a nationally representative sample of individuals across adolescence and into early adulthood we found that bullying and victimisation in adolescence were associated with higher proportions of sexual minority individuals reporting adverse mental health at age 23 compared to heterosexual individuals with the same frequency of bullying/victimisation. We confirm previous findings of higher rates of bullying experiences in sexual minority individuals across adolescence in the UK. The limited longitudinal studies in the UK have found that the higher rates of mental health problems reported by sexual minority adolescents is explained to a greater extent by their higher levels of bullying compared to heterosexual peers^23 24^. Another UK study also using data from the MCS found that sexual minority adolescents were more likely to belong to classes with higher levels of victimisation which partly explained mental health inequities^23^. However, these studies were limited to adolescence only, did not examine the role of loneliness and lack of social support, and used different statistical methods due to differing research aims.

We also found that there was a gradient with proportions of individuals reporting adverse mental health increasing with the frequency of bullying/victimisation, i.e., from never experiencing bullying/victimisation, experiencing it once and experiencing it at ≥two time points. A final key finding from this study was the differential role that loneliness and social support played for sexual minority youth compared to heterosexual youth. Findings show that experiencing loneliness and lack of peer and family support explained 15% to 45% of the predicted probabilities of adverse mental health outcomes in sexual minority groups but not in heterosexual individuals.

The largely consistent, non-significant interaction terms between sexual identity and bullying/victimisation across all mental health outcomes imply that bullying/victimisation is strongly associated with poor mental health, regardless of sexual identity. However, our findings highlight that the massive health inequities observed in the predictive probabilities for all outcomes are driven by the compounding double disadvantage of sexual minority identity intersectionally layered with cumulative exposure to bullying/victimisation in adolescence.

### Strengths and limitations

Key strengths include a large and nationally representative sample with assessments of bullying and victimisation at multiple points between ages 11-23. Consistent associations with similar patterns were found for all mental health outcomes in relation to bullying and victimisation, reenforcing the deleterious and long-term impact on mental health. The questions on bullying and victimisation in the MCS capture different aspects including traditional bullying by peers and siblings both in-person and online and being victimised for a wide range of reasons. The relatively large sample size of sexual minority individuals enabled examining differences by sexual minority sub-group often overlooked in studies. The longitudinal design with bullying and victimisation recorded before mental health outcomes is a key strength. It is a common strategy to prospectively collect data across adolescence, sexual identity in late adolescence and link it back to early life exposures^24^.

Like most cohort studies, the MCS suffers from attrition with significant loss to follow-up over 23 years. However, studies indicate that the sample at ages 17 and 23 remain representative of the original birth cohort^31^. We addressed missing data using multiple imputation with a rich dataset including several auxiliary variables further supported by longitudinal data assessed across childhood greatly increasing the validity and robustness of imputation, i.e., auxiliary variables help predict missing data with greater precision and minimising non-random variation in the values^32^. Some individuals may not recall distressing past events for bullying/victimisation and self-harm and attempted suicide leading to an underestimation of associations. We were unable to examine specific forms of bullying including being bullied or victimised for sexual minority identity/appearance or separate online from in-person bullying or the different forms of victimisation due to small numbers or data limitations. We were also unable to examine whether associations differed by ethnicity and/or childhood socioeconomic circumstances (i.e., using interaction terms) due to the smaller numbers of individuals identifying with both minority identities. Sexual identity was measured at age 17 and some individuals may change their sexuality across adolescence which we could not account for. These limitations must be addressed in the future studies which are adequately powered and with wider range of information. Lastly, bullying and victimisation were not assessed at all five ages. While questions were framed nearly identically across follow-up there are some differences like online bullying being recorded at only age 14 reflecting common issues and changing priorities as cohorts progress over time. We used validated measures of loneliness and social support, but the MCS does not collect information on sexual minority relevant issues like number of people who are aware of the participants’ sexual minority identity and both in-person and online access to sexual minority communities which can moderate the impact of bullying on mental health.

### Implications for policy and research

The UK government recognises the long-term deleterious impact of bullying on mental health and wellbeing, and mandates schools implement procedures to prevent various forms of bullying, including cyber- and in-person bullying. Guidance explicitly requires the proactive prevention of victimisation based on protected characteristics, such as sexual orientation, gender, ethnicity and disability. Further, schools have the authority to prevent bullying off-premises and are meant to devise their own anti-bullying policies and can be held accountable for their failure to prevent bullying. Despite the comprehensive guidance and laws to prevent bullying, national surveys demonstrate that bullying especially among adolescents remain substantially high. Current research on the efficacy of these school-level policies is limited and often hindered by selection bias, as studies frequently rely on regional samples or participation of schools with established best practices. These limited studies indicate increase in policy coverage in recent years including bias- and homophobic bullying^13^. Further, having strong policies to tackle bullying does not necessarily guarantee effective implementation.

While eliminating bullying entirely may be an elusive goal, stronger policies are required, more consistent enforcement, and rigorous auditing of school policies. Further, national surveys on bullying experiences in children should oversample sexual minority individuals to better track the prevalence and change in identity-based bullying. Still, substantially more can be done to empower sexual minority adolescents both in schools and environments like public transport, neighbourhoods and community centres. For example, schools can take proactive steps to employ staff members who identify as sexual minority, including counsellors who can provide much needed guidance and support. Similarly, staff can be employed in public transport, neighbourhoods, shopping malls and other areas frequented by adolescents to help prevent and sensitise the public on bullying, and importantly such measures need to be encouraged across the country going beyond large urban areas. Adopting bystander-focused models like the Finnish "KiVa" program—which has proven effective in reducing victimisation—could be particularly beneficial for marginalized groups who may otherwise fear the repercussions of reporting^14^. Our pertinent findings on the role of loneliness and social support in mental health inequalities highlight the need for constructive in-person and online support for sexual minority adolescents. This could include proactive digital engagement to access moderated LGBTQ+ communities, easier access to LGBTQ+ organisations, and establishing inter-school LGBTQ+ organisations. However, reducing loneliness and lack of social support will require changes to the broader environment including in schools and at homes. For example, staff can be trained on inclusive policies like LGBTQ+ affirming behaviours, family-based support to understand the impact of and addressing stressors like bullying and building trusting environments at home and elsewhere.

### Final summary

Sexual minority individuals including those who identify as mainly heterosexual, bisexual, gay/lesbian and other sexual minority identities report 2-3 times higher risk for cumulative experiences of bullying and victimisation across adolescence and into early adulthood. Experiencing cumulative bullying and victimisation is associated with higher risk for poor mental health in sexual minority groups compared to heterosexual peers experiencing the same level of bullying or victimisation. Loneliness and lack of social support explain a substantial portion of the higher risk for poor mental health in sexual minority but not heterosexual groups. Better and more effective policies are urgently required to reduce bullying and victimisation.

## Data Availability

The data used in this study is from the Millennium Cohort Study and the data can be downloaded via special license and after registering on the UK Data Service website (https://ukdataservice.ac.uk/) and agreeing to all terms and conditions.

