## Supplemental Tables for "Sexual orientation inequalities in mental health across adolescence and early adulthood: exploring the contribution of cumulative experiences of bullying and victimisation over time"

**Supplemental Table 1. A detailed description of key variables including exposures, mental health indicators and covariates used in this study**

| Variables of interest used in this study | Question(s) in cohort member computer-assisted personal interview (CAPI), self-completion interview (CASI) or online questionnaire (CAWI) | Final version used in analysis | Comments |
|---|---|---|---|
| **<u>Bullying and victimisation</u>** | | | |
| Bullying based on three questions at ages 11 & 14 | | | |
| | At ages 11 & 14:<br>'How often do other children hurt you/pick on you on purpose?'<br><br>"How often do your brothers or sisters hurt you or pick on you on purpose?"<br><br>Age 14 only:<br>"How often do other children bully you online" | Binary:<br>most days/weekly indicating frequent bullying vs. monthly/every few months/less often/never indicating less frequent/never bullied<br><br>Binary:<br>most days/weekly indicating frequent bullying vs. monthly/every few months/less often/never indicating less frequent/never bullied<br><br>A final categorical variable was created to indicate bullying frequency in early adolescence:<br>0: never bullied<br>1: bullied once<br>2: Bullied 2 times | |
| Victimisation at ages 14, 17 & 23 | "In the past 12 months has anyone done any of these things to you?"<br><br>Options:<br>1. Insulted/threatened/shouted, 2. Experiences of physical violence, 3. Hit or used a weapon, 4. Had something stolen and 5. Been sexually assaulted. | Binary variable at each age:<br><br>1: No experiences of victimisation<br>2: Any experience of any kind of victimisation | |

| | | | |
|---|---|---|---|
| | Response options for each question:<br>1. Yes<br>2. No | A final categorical variable was created to indicate victimisation frequency:<br>0: never victimised<br>1: victimised once<br>2: victimised 2/3 times | |
| **<u>Mental health at age 23</u>** | | | |
| Self-reported Kessler (6 item) | During the last 30 days about how often:<br>- did you feel so depressed that nothing could cheer you up?<br>- did you feel hopeless?<br>- did you feel restless or fidgety?<br>- did you feel that everything was an effort?<br>- did you feel worthless?<br>- did you feel nervous? | Binary:<br><br>1: 0-12: No psychological distress<br>2: 13-24: indicates psychological distress (symptoms of depression and anxiety) | Each item coded 0-4 add total summed (range 0 to 24)<br><br>4. All of the time<br>3. Most of the time<br>2. Some of the time<br>1. A little of the time<br>0. None of the time |
| Self-harm | During the last year, have you hurt yourself on purpose in any of the following ways?<br>Cut or stabbed yourself<br>Burned yourself<br>Bruised or pinched yourself<br>Taken an overdose of tablets<br>Pulled out your hair<br>Hurt yourself some other way | Binary:<br><br>No vs yes (any kind of self-harm) | For each self-harming action, options included:<br>1. Yes<br>2. No |
| Attempted suicide | Have you ever hurt yourself on purpose in an attempt to end your life? | Binary:<br>No vs yes | |
| UCL Loneliness Scale | How often do you feel that you lack companionship?<br>How often do you feel left out?<br>How often do you feel isolated from others? | Binary:<br><br>1. 3-5: "not lonely"<br>2. 6-9: "lonely" | For each question options include:<br>1 Hardly ever<br>2 Some of the time<br>3 Often<br>Final summary score ranges from 3 to 9 |
| Social support | In answering the following questions, think about your current relationships with friends, family members, community members, and so on. Please indicate to what | Binary:<br><br>1. Very true/partly true<br>2. Not true at all | Participants could choose one of three possible answers:<br>1 Very true<br>2 Partly true |

| | | | |
|---|---|---|---|
| | extent each statement describes your current relationships with other people.<br><br>"I have family and friends who help me feel safe, secure and happy." | | 3 Not true at all |
| Sexual identity at age 17 | Which of the following options best describes how you currently think of yourself?<br>1 Completely heterosexual / straight<br>2 Mainly heterosexual / straight<br>3 Bisexual<br>4 Mainly gay or lesbian<br>5 Completely gay or lesbian<br>6 Other<br>7 Don't know<br>8 Prefer not to say | Categorical:<br><br>1 Completely heterosexual / straight<br>2 Mainly heterosexual / straight<br>3 Bisexual<br>4 Gay or lesbian<br>5 Other sexual identity groups | |

**Supplemental Table 2. Comparison of complete case (CC) and multiply imputed (MI) data**

| | CC% [95% CI] | MI % [95% CI] |
|---|---|---|
| **Bullying, age 11** | | |
| No | 54.1 [52.5- 55.7] | 55.7 [54.5- 56.8] |
| Yes | 45.9 [44.3- 47.5] | 44.3 [43.2- 45.5] |
| **Bullying, age 14** | | |
| No | 68.2 [66.7- 69.7] | 69.4 [68.3- 70.5] |
| Yes | 31.8 [30.3- 33.3] | 30.6 [29.5- 31.7] |
| **Victimisation, age 14** | | |
| No | 49.5 [47.9- 51.1] | 51.4 [50.4- 52.4] |
| Yes | 50.5 [48.9- 52.1] | 48.6 [47.6- 49.6] |
| **Victimisation, age 17** | | |
| No | 51.9 [50.3- 53.5] | 54.9 [54.0- 55.8] |
| Yes | 48.1 [46.5- 49.7] | 45.1 [44.2- 46.0] |
| **Victimisation, age 23** | | |
| No | 70.7 [69.2- 72.1] | 75.1 [74.3- 76.0] |
| Yes | 29.3 [27.9- 30.8] | 24.9 [24.0- 25.7] |
| **Self-harm, age 23** | | |
| No | 77.4 [76.0- 78.7] | 79.5 [78.7- 80.4] |
| Yes | 22.6 [21.3- 24.0] | 20.5 [19.6- 21.3] |
| **Attempted suicide** | | |
| No | 90.0 [89.0- 90.9] | 89.4 [88.8- 90.1] |
| Yes | 10.0 [ 9.1- 11.0] | 10.6 [ 9.9- 11.2] |
| **Sexual identity** | | |
| Heterosexual | 75.7 [74.3- 77.0] | 78.4 [77.6-79.2] |
| Mainly heterosexual | 13.3 [12.2- 14.4] | 10.5 [9.9- 11.1] |
| Bisexual | 7.6 [ 6.8-8.5] | 6.4 [5.9-6.9] |
| Gay/lesbian | 2.6 [ 2.1-3.1] | 2.6 [2.2-2.9] |
| OSI | 0.9 [ 0.7-1.3] | 2.1 [1.8-2.4] |
| **Loneliness** | | |
| | 84.1 [82.9-85.3] | 82.8 [82.0-83.6] |
| | 15.9 [14.7-17.1] | 17.2 [16.4-18.0] |

| | | |
|---|---|---|
| **Social support** | | |
| | 83.4 [82.1-84.5] | 79.8 [78.9-80.7] |
| | 16.6 [ 15.5-17.9] | 20.2 [ 19.3-21.1] |
| | **Mean (SE)** | **Mean (SE)** |
| **Kessler 6** | 8.2 (4.9) | 8.4 [0.1] |
| **BMI, age 11** | 18.8 (3.2) | 19.3 [0.0] |
| **BMI, age 14** | 21.1 (3.8) | 21.6 [0.0] |
| **BMI, age 17** | 23.0 (4.4) | 23.5 [0.0] |
| **BMI, age 23** | 25.1 (4.9) | 25.6 [0.1] |

**Supplemental Table 3. Associations between sexual identity and experiences of cumulative bullying and victimisation across adolescence in 12,782 individuals from the Millennium Cohort Study**

| | **Bullying** | | | | **Victimisation** | | | |
|---|---|---|---|---|---|---|---|---|
| | **Model 1** | | **Model 2** | | **Model 1** | | **Model 2** | |
| | **RRR** | **95% CI** | **RRR** | **95% CI** | **RRR** | **95% CI** | **RRR** | **95% CI** |
| ***Never bullied/victimised*** | Ref | | Ref | | Ref | | Ref | |
| **1 time** | | | | | | | | |
| Heterosexual | 1 | | 1 | | 1 | | 1 | |
| Mainly heterosexual | 1.10 | 0.93,1.31 | 1.12 | 0.94,1.33 | **1.48** | **1.19,1.84** | **1.50** | **1.20,1.86** |
| Bisexual | **1.29** | **1.05,1.60** | **1.29** | **1.05,1.60** | **1.38** | **1.03,1.85** | **1.40** | **1.05,1.87** |
| Gay/lesbian | 1.34 | 0.93,1.92 | 1.32 | 0.92,1.91 | 1.36 | 0.87,2.13 | 1.33 | 0.85,2.09 |
| Other | **1.59** | **1.08,2.32** | **1.55** | **1.05,2.28** | 1.10 | 0.71,1.70 | 1.12 | 0.73,1.73 |
| sex | | | 1.01 | 0.91,1.11 | | | 0.92 | 0.82,1.04 |
| **Ethnicity** | | | | | | | | |
| White | | | 1 | | | | 1 | |
| Non-White | | | 0.89 | 0.77,1.03 | | | 0.77 | 0.66,0.89 |
| **Parental income** | | | | | | | | |
| Lowest quintile | | | 1 | | | | 1 | |
| Q2 | | | 1.02 | 0.84,1.22 | | | 1.20 | 0.99,1.46 |
| Q3 | | | 0.83 | 0.69,0.99 | | | 1.04 | 0.86,1.27 |
| Q4 | | | 0.82 | 0.69,0.97 | | | 1.12 | 0.92,1.37 |
| Highest quintile | | | 0.77 | 0.65,0.92 | | | 1.06 | 0.89,1.28 |
| **≥2 times** | | | | | | | | |
| Heterosexual | 1 | | 1 | | 1 | | 1 | |
| Mainly heterosexual | **1.45** | **1.17,1.79** | **1.45** | **1.17,1.81** | **2.39** | **1.93,2.96** | **2.43** | **1.95,3.02** |
| Bisexual | **1.70** | **1.37,2.12** | **1.64** | **1.31,2.05** | **3.13** | **2.41,4.07** | **3.18** | **2.44,4.15** |
| Gay/lesbian | **2.72** | **1.87,3.95** | **2.64** | **1.81,3.86** | **2.76** | **1.85,4.13** | **2.68** | **1.79,4.00** |
| Other | **2.61** | **1.73,3.94** | **2.45** | **1.62,3.70** | 1.41 | 0.95,2.10 | 1.45 | 0.98,2.17 |
| sex | | | **1.16** | **1.03,1.32** | | | **0.88** | **0.78,0.99** |
| **Ethnicity** | | | | | | | | |
| White | | | 1 | | | | 1 | |

| | | | | | | | | |
|---|---|---|---|---|---|---|---|---|
| Non-White | | | 0.83 | 0.69,1.00 | | | 0.62 | 0.53,0.72 |
| **Parental income** | | | | | | | | |
| Lowest quintile | | | 1 | | | | 1 | |
| Q2 | | | 1.37 | 1.11,1.70 | | | **1.38** | **1.12,1.71** |
| Q3 | | | 1.00 | 0.81,1.23 | | | 1.23 | 1.00,1.50 |
| Q4 | | | 0.87 | 0.70,1.10 | | | 1.20 | 0.97,1.48 |
| Highest quintile | | | **0.74** | **0.58,0.94** | | | 1.13 | 0.92,1.38 |

Model 1: Unadjusted, Model 2: adjusted for assigned sex at birth, ethnicity and parental income. Text in bold indicates RRR with 95% CIs that do not include 1.

**Supplemental Table 4. Associations between frequency of bullying in adolescence and mental health outcomes in early adulthood in 12,782 individuals from the Millennium Cohort Study**

| | Psychological distress | | | | | | | | Self-harm | | | | | | | | Attempted suicide | | | | | | | |
|---|---|---|---|---|---|---|---|---|---|---|---|---|---|---|---|---|---|---|---|---|---|---|---|---|
| | Model 1 | | Model 2 | | Model 3 | | Model 4 | | Model 1 | | Model 2 | | Model 3 | | Model 4 | | Model 1 | | Model 2 | | Model 3 | | Model 4 | |
| **Bullying** | **OR** | **95% CI** | **OR** | **95% CI** | **OR** | **95% CI** | **OR** | **95% CI** | **OR** | **95% CI** | **OR** | **95% CI** | **OR** | **95% CI** | **OR** | **95% CI** | **OR** | **95% CI** | **OR** | **95% CI** | **OR** | **95% CI** | **OR** | **95% CI** |
| None | 1 | | 1 | | 1 | | 1 | | 1 | | 1 | | 1 | | 1 | | 1 | | 1 | | 1 | | 1 | |
| 1 time | 1.34 | 1.13,1.60 | 1.32 | 1.10,1.57 | 1.25 | 1.03,1.51 | 1.2 | 0.99,1.47 | 1.16 | 0.98,1.37 | 1.15 | 0.97,1.36 | 1.1 | 0.93,1.31 | 1.07 | 0.90,1.28 | 1.25 | 0.97,1.60 | 1.22 | 0.95,1.56 | 1.16 | 0.90,1.50 | 1.13 | 0.87,1.45 |
| 2 times | 1.89 | 1.57,2.28 | 1.8 | 1.49,2.17 | 1.56 | 1.28,1.91 | 1.51 | 1.23,1.86 | 1.54 | 1.25,1.90 | 1.5 | 1.21,1.85 | 1.35 | 1.08,1.69 | 1.31 | 1.04,1.64 | 1.92 | 1.49,2.48 | 1.8 | 1.39,2.33 | 1.63 | 1.25,2.13 | 1.57 | 1.20,2.07 |
| **Sexual identity** | | | | | | | | | | | | | | | | | | | | | | | | |
| Heterosexual | 1 | | 1 | | 1 | | 1 | | 1 | | 1 | | 1 | | 1 | | 1 | | 1 | | 1 | | 1 | |
| Mainly heterosexual | 1.81 | 1.34,2.45 | 1.81 | 1.34,2.44 | 1.52 | 1.10,2.09 | 1.43 | 1.04,1.98 | 2.01 | 1.49,2.72 | 1.91 | 1.41,2.58 | 1.71 | 1.25,2.32 | 1.64 | 1.20,2.24 | 1.49 | 0.98,2.28 | 1.46 | 0.96,2.22 | 1.3 | 0.85,1.97 | 1.22 | 0.80,1.88 |
| Bisexual | 3.46 | 2.44,4.90 | 3.36 | 2.36,4.79 | 2.7 | 1.83,3.99 | 2.51 | 1.67,3.76 | 4.76 | 3.46,6.54 | 4.35 | 3.16,6.00 | 3.77 | 2.72,5.22 | 3.58 | 2.57,4.98 | 3.92 | 2.54,6.04 | 3.7 | 2.38,5.74 | 3.15 | 2.02,4.90 | 2.91 | 1.85,4.58 |
| Gay/lesbian | 2.78 | 1.58,4.91 | 2.72 | 1.53,4.82 | 2.51 | 1.32,4.76 | 2.4 | 1.25,4.59 | 3.56 | 1.88,6.76 | 3.33 | 1.75,6.33 | 3.11 | 1.57,6.14 | 3.02 | 1.51,6.03 | 4.17 | 2.15,8.08 | 3.95 | 2.00,7.82 | 3.67 | 1.84,7.31 | 3.54 | 1.75,7.16 |
| other | 4.08 | 2.24,7.44 | 3.75 | 1.98,7.11 | 2.67 | 1.31,5.44 | 2.17 | 1.02,4.63 | 3.24 | 1.72,6.08 | 3.02 | 1.60,5.69 | 2.32 | 1.18,4.56 | 1.96 | 0.98,3.93 | 4.3 | 2.05,9.03 | 3.86 | 1.81,8.22 | 3.04 | 1.40,6.61 | 2.53 | 1.18,5.42 |
| **Interactions between bullying & sexual identity** | | | | | | | | | | | | | | | | | | | | | | | | |
| bully_total=0 ~o | 1 | | 1 | | 1 | | 1 | | 1 | | 1 | | 1 | | 1 | | 1 | | 1 | | 1 | | 1 | |
| bully_total=1 ~o | 1.06 | 0.71,1.60 | 1.07 | 0.71,1.61 | 1.04 | 0.67,1.62 | 1.09 | 0.70,1.71 | 1.32 | 0.87,2.03 | 1.35 | 0.88,2.06 | 1.33 | 0.86,2.06 | 1.39 | 0.88,2.18 | 1.46 | 0.85,2.50 | 1.47 | 0.85,2.54 | 1.46 | 0.84,2.53 | 1.52 | 0.87,2.68 |
| bully_total=1 ~l | 0.88 | 0.52,1.48 | 0.83 | 0.49,1.40 | 0.94 | 0.53,1.67 | 0.96 | 0.53,1.74 | 0.98 | 0.60,1.58 | 0.96 | 0.59,1.56 | 1.04 | 0.63,1.72 | 1.06 | 0.64,1.75 | 1.71 | 0.97,3.01 | 1.64 | 0.93,2.91 | 1.8 | 1.01,3.20 | 1.88 | 1.04,3.40 |
| bully_total=1 ~b | 1.02 | 0.47,2.24 | 1.07 | 0.49,2.36 | 0.97 | 0.40,2.34 | 0.94 | 0.38,2.35 | 0.78 | 0.33,1.85 | 0.79 | 0.33,1.90 | 0.73 | 0.29,1.83 | 0.7 | 0.27,1.81 | 1.02 | 0.42,2.47 | 1.06 | 0.43,2.62 | 1 | 0.40,2.51 | 0.96 | 0.37,2.49 |
| bully_total=1 ~r | 1.06 | 0.45,2.52 | 1.03 | 0.41,2.58 | 1.18 | 0.43,3.26 | 1.32 | 0.46,3.77 | 0.88 | 0.38,2.02 | 0.87 | 0.38,1.99 | 0.92 | 0.38,2.22 | 0.99 | 0.39,2.46 | 1.11 | 0.43,2.89 | 1.08 | 0.41,2.86 | 1.16 | 0.42,3.17 | 1.27 | 0.45,3.56 |
| bully_total=2 ~o | 1 | .,. | 1 | | 1 | | 1 | | 1 | | 1 | | 1 | | 1 | | 1 | | 1 | | 1 | | 1 | |
| bully_total=2 ~o | 1.07 | 0.70,1.62 | 1.09 | 0.71,1.66 | 1.21 | 0.77,1.92 | 1.22 | 0.76,1.96 | 1.25 | 0.80,1.93 | 1.28 | 0.82,1.98 | 1.36 | 0.86,2.15 | 1.37 | 0.86,2.18 | 1.36 | 0.75,2.47 | 1.4 | 0.77,2.54 | 1.48 | 0.81,2.71 | 1.49 | 0.80,2.76 |
| bully_total=2 ~l | 0.79 | 0.46,1.37 | 0.73 | 0.42,1.28 | 0.73 | 0.41,1.31 | 0.74 | 0.40,1.35 | 0.8 | 0.47,1.37 | 0.79 | 0.46,1.35 | 0.79 | 0.45,1.38 | 0.79 | 0.45,1.40 | 0.92 | 0.51,1.68 | 0.85 | 0.46,1.58 | 0.87 | 0.47,1.61 | 0.87 | 0.47,1.64 |
| bully_total=2 ~b | 1.05 | 0.50,2.22 | 1.09 | 0.52,2.31 | 1.01 | 0.45,2.27 | 1.02 | 0.44,2.34 | 1.02 | 0.46,2.30 | 1.09 | 0.49,2.44 | 1.03 | 0.44,2.43 | 1.03 | 0.43,2.47 | 0.85 | 0.36,2.03 | 0.89 | 0.37,2.17 | 0.84 | 0.34,2.08 | 0.83 | 0.32,2.13 |

| | | | | | | | | | | | | | | | | | | | | | | | | |
|---|---|---|---|---|---|---|---|---|---|---|---|---|---|---|---|---|---|---|---|---|---|---|---|---|
| bully_total=2 ~r | 1.07 | 0.42,2.69 | 1.03 | 0.40,2.65 | 0.92 | 0.33,2.58 | 0.98 | 0.34,2.83 | 0.98 | 0.42,2.31 | 0.94 | 0.40,2.19 | 0.87 | 0.36,2.10 | 0.9 | 0.36,2.21 | 1.16 | 0.43,3.13 | 1.13 | 0.41,3.10 | 1.08 | 0.38,3.06 | 1.14 | 0.40,3.21 |
| **Sex** | | | | | | | | | | | | | | | | | | | | | | | | |
| Female | | | 1.59 | 1.42,1.78 | 1.63 | 1.44,1.84 | 1.77 | 1.56,2.00 | | | 1.52 | 1.34,1.71 | 1.52 | 1.34,1.72 | 1.6 | 1.41,1.82 | | | 1.63 | 1.38,1.93 | 1.62 | 1.36,1.92 | 1.72 | 1.45,2.05 |
| **Ethnicity** | | | | | | | | | | | | | | | | | | | | | | | | |
| White | | | 1 | | 1 | | 1 | | | | 1 | | 1 | | 1 | | | | 1 | | 1 | | 1 | |
| Non-White | | | 1.06 | 0.90,1.25 | 1.04 | 0.88,1.24 | 0.98 | 0.82,1.17 | | | 0.8 | 0.67,0.96 | 0.78 | 0.65,0.94 | 0.73 | 0.61,0.89 | | | 0.79 | 0.61,1.03 | 0.78 | 0.60,1.01 | 0.73 | 0.56,0.94 |
| **Parental income** | | | | | | | | | | | | | | | | | | | | | | | | |
| Lower quintile | | | 1 | | 1 | | 1 | | | | 1 | | 1 | | 1 | | | | 1 | | 1 | | 1 | |
| Q2 | | | 0.97 | 0.80,1.18 | 1.01 | 0.81,1.25 | 1.03 | 0.82,1.29 | | | 0.92 | 0.73,1.16 | 0.94 | 0.74,1.19 | 0.96 | 0.75,1.22 | | | 0.95 | 0.71,1.27 | 0.96 | 0.71,1.30 | 0.98 | 0.72,1.34 |
| Q3 | | | 0.77 | 0.62,0.96 | 0.83 | 0.66,1.04 | 0.9 | 0.71,1.15 | | | 0.87 | 0.70,1.09 | 0.92 | 0.73,1.17 | 1 | 0.79,1.28 | | | 0.75 | 0.57,0.98 | 0.78 | 0.59,1.04 | 0.86 | 0.64,1.16 |
| Q4 | | | 0.61 | 0.50,0.74 | 0.64 | 0.52,0.80 | 0.73 | 0.59,0.91 | | | 0.89 | 0.73,1.10 | 0.96 | 0.77,1.19 | 1.08 | 0.87,1.35 | | | 0.54 | 0.41,0.70 | 0.57 | 0.43,0.74 | 0.64 | 0.49,0.85 |
| Highest quintile | | | 0.47 | 0.39,0.57 | 0.49 | 0.40,0.60 | 0.58 | 0.46,0.72 | | | 0.74 | 0.61,0.90 | 0.8 | 0.65,0.98 | 0.93 | 0.76,1.15 | | | 0.46 | 0.35,0.61 | 0.49 | 0.37,0.65 | 0.58 | 0.43,0.78 |
| **Loneliness** | | | | | | | | | | | | | | | | | | | | | | | | |
| Not lonely | | | | | 1 | | 1 | | | | | | 1 | | 1 | | | | | | 1 | | 1 | |
| lonely | | | | | 6.35 | 5.59,7.22 | 4.76 | 4.17,5.44 | | | | | 3.12 | 2.76,3.53 | 2.4 | 2.11,2.73 | | | | | 2.7 | 2.28,3.20 | 1.95 | 1.63,2.35 |
| **Support** | | | | | | | | | | | | | | | | | | | | | | | | |
| Yes | | | | | | | 1 | | | | | | | | 1 | | | | | | | | 1 | |
| No | | | | | | | 3.1 | 2.67,3.60 | | | | | | | 2.56 | 2.21,2.98 | | | | | | | 2.82 | 2.32,3.43 |

Model 1 (crude, unadjusted), Model 2: adjusted for ASAB, ethnicity, parental income, Model 3: additionally adjusted for loneliness, and Model 4: additionally adjusted for support. All models included interaction terms between sexual identity and bullying.

**Supplemental Table 5. Associations between frequency of victimisation in adolescence and mental health outcomes in early adulthood in 12,782 individuals from the Millennium Cohort Study**

| | Psychological distress | | | | | | | | Self-harm | | | | | | | | Attempted suicide | | | | | | | |
|---|---|---|---|---|---|---|---|---|---|---|---|---|---|---|---|---|---|---|---|---|---|---|---|---|
| | Model 1 | | Model 2 | | Model 3 | | Model 4 | | Model 1 | | Model 2 | | Model 3 | | Model 4 | | Model 1 | | Model 2 | | Model 3 | | Model 4 | |
| **Victimisation** | **OR** | **95% CI** | **OR** | **95% CI** | **OR** | **95% CI** | **OR** | **95% CI** | **OR** | **95% CI** | **OR** | **95% CI** | **OR** | **95% CI** | **OR** | **95% CI** | **OR** | **95% CI** | **OR** | **95% CI** | **OR** | **95% CI** | **OR** | **95% CI** |
| None | 1 | | 1 | | 1 | | 1 | | 1 | | 1 | | 1 | | 1 | | 1 | | 1 | | 1 | | 1 | |
| 1 time | 1.51 | 1.22,1.86 | 1.56 | 1.26,1.93 | 1.35 | 1.08,1.68 | 1.31 | 1.05,1.64 | 1.78 | 1.44,2.21 | 1.8 | 1.45,2.23 | 1.64 | 1.32,2.05 | 1.62 | 1.29,2.02 | 2.02 | 1.46,2.81 | 2.06 | 1.48,2.87 | 1.9 | 1.36,2.65 | 1.85 | 1.33,2.58 |
| 2 times | 2.6 | 2.19,3.09 | 2.75 | 2.31,3.28 | 2.19 | 1.82,2.62 | 2.0 | 1.66,2.41 | 3.12 | 2.56,3.80 | 3.18 | 2.60,3.89 | 2.74 | 2.22,3.37 | 2.57 | 2.08,3.17 | 4.29 | 3.17,5.82 | 4.43 | 3.25,6.04 | 3.83 | 2.80,5.23 | 3.52 | 2.57,4.82 |
| **Sexual identity** | | | | | | | | | | | | | | | | | | | | | | | | |
| Heterosexual | 1 | | 1 | | 1 | | 1 | | 1 | | 1 | | 1 | | 1 | | 1 | | 1 | | 1 | | 1 | |
| Mainly heterosexual | 1.39 | 0.85,2.26 | 1.42 | 0.87,2.32 | 1.1 | 0.65,1.88 | 1.05 | 0.63,1.77 | 2.57 | 1.66,3.98 | 2.49 | 1.61,3.84 | 2.19 | 1.42,3.39 | 2.14 | 1.37,3.35 | 2 | 0.96,4.17 | 2.01 | 0.97,4.17 | 1.77 | 0.84,3.70 | 1.71 | 0.81,3.61 |
| Bisexual | 2.76 | 1.61,4.73 | 2.62 | 1.53,4.49 | 1.93 | 1.08,3.43 | 1.77 | 0.97,3.22 | 4.34 | 2.62,7.17 | 3.96 | 2.37,6.62 | 3.3 | 1.94,5.61 | 3.15 | 1.81,5.48 | 2.29 | 0.88,5.95 | 2.07 | 0.79,5.44 | 1.73 | 0.66,4.55 | 1.55 | 0.58,4.15 |
| Gay/lesbian | 2.96 | 1.31,6.69 | 3.08 | 1.35,7.05 | 2.78 | 1.16,6.67 | 2.69 | 1.09,6.59 | 4.68 | 2.15,10.17 | 4.33 | 2.01,9.33 | 3.99 | 1.78,8.95 | 3.91 | 1.70,9.02 | 5.5 | 2.01,15.03 | 5.42 | 1.93,15.22 | 5 | 1.82,13.74 | 4.86 | 1.72,13.75 |
| other | 4.4 | 2.31,8.38 | 3.65 | 1.89,7.03 | 2.84 | 1.39,5.82 | 2.54 | 1.23,5.25 | 4.27 | 2.18,8.38 | 3.88 | 2.00,7.56 | 3.2 | 1.62,6.33 | 2.92 | 1.48,5.78 | 4.58 | 1.75,11.96 | 3.78 | 1.46,9.80 | 3.2 | 1.18,8.64 | 2.87 | 1.03,7.97 |
| **Interactions between victimisation & sexual identity** | | | | | | | | | | | | | | | | | | | | | | | | |
| vic_total2=0 #~o | 1 | | 1 | | 1 | | 1 | | 1 | | 1 | | 1 | | 1 | | 1 | | 1 | | 1 | | 1 | |
| vic_total2=1 #~o | 1.29 | 0.71,2.34 | 1.23 | 0.67,2.25 | 1.44 | 0.76,2.72 | 1.42 | 0.76,2.65 | 0.69 | 0.40,1.19 | 0.68 | 0.39,1.18 | 0.72 | 0.41,1.24 | 0.7 | 0.40,1.22 | 0.68 | 0.27,1.71 | 0.65 | 0.26,1.64 | 0.69 | 0.27,1.75 | 0.67 | 0.26,1.69 |
| vic_total2=1 #~l | 0.8 | 0.39,1.64 | 0.79 | 0.39,1.61 | 0.93 | 0.44,1.97 | 0.95 | 0.43,2.09 | 0.95 | 0.49,1.84 | 0.95 | 0.48,1.86 | 1.06 | 0.52,2.12 | 1.07 | 0.52,2.20 | 1.71 | 0.57,5.14 | 1.73 | 0.57,5.30 | 1.9 | 0.61,5.95 | 2.02 | 0.64,6.43 |
| vic_total2=1 #~b | 0.74 | 0.24,2.28 | 0.74 | 0.23,2.33 | 0.7 | 0.22,2.27 | 0.68 | 0.20,2.32 | 0.4 | 0.14,1.17 | 0.43 | 0.15,1.26 | 0.41 | 0.14,1.24 | 0.4 | 0.13,1.25 | 0.55 | 0.15,1.99 | 0.56 | 0.15,2.10 | 0.54 | 0.15,2.02 | 0.52 | 0.14,1.97 |
| vic_total2=1 #~r | 0.86 | 0.35,2.12 | 0.91 | 0.36,2.30 | 0.85 | 0.31,2.37 | 0.86 | 0.30,2.46 | 0.44 | 0.16,1.16 | 0.43 | 0.16,1.15 | 0.41 | 0.15,1.10 | 0.4 | 0.15,1.07 | 0.71 | 0.21,2.45 | 0.71 | 0.21,2.47 | 0.69 | 0.19,2.50 | 0.68 | 0.18,2.57 |
| vic_total2=2 #~o | 1 | | 1 | | 1 | | 1 | | 1 | | 1 | | 1 | | 1 | | 1 | | 1 | | 1 | | 1 | |
| vic_total2=2 #~o | 1.27 | 0.75,2.16 | 1.25 | 0.74,2.11 | 1.44 | 0.81,2.56 | 1.5 | 0.84,2.67 | 0.85 | 0.52,1.38 | 0.84 | 0.52,1.37 | 0.89 | 0.54,1.46 | 0.91 | 0.55,1.51 | 0.85 | 0.38,1.89 | 0.84 | 0.38,1.85 | 0.88 | 0.39,1.97 | 0.9 | 0.40,2.02 |
| vic_total2=2 #~l | 1.06 | 0.59,1.90 | 1.02 | 0.56,1.84 | 1.27 | 0.67,2.40 | 1.33 | 0.69,2.56 | 0.86 | 0.48,1.53 | 0.83 | 0.46,1.50 | 0.93 | 0.50,1.73 | 0.95 | 0.50,1.79 | 1.87 | 0.70,5.00 | 1.86 | 0.69,4.95 | 2.07 | 0.77,5.56 | 2.24 | 0.82,6.10 |

| | | | | | | | | | | | | | | | | | | | | | | | | |
|---|---|---|---|---|---|---|---|---|---|---|---|---|---|---|---|---|---|---|---|---|---|---|---|---|
| vic_total 2=2 #~b | 0.97 | 0.39,2.44 | 0.9 | 0.35,2.30 | 0.88 | 0.32,2.44 | 0.88 | 0.31,2.50 | 0.69 | 0.30,1.61 | 0.71 | 0.31,1.63 | 0.7 | 0.29,1.68 | 0.69 | 0.28,1.71 | 0.64 | 0.22,1.87 | 0.62 | 0.21,1.86 | 0.61 | 0.21,1.78 | 0.6 | 0.20,1.84 |
| vic_total 2=2 #~r | 1.2 | 0.53,2.72 | 1.35 | 0.58,3.13 | 1.17 | 0.46,2.94 | 1.1 | 0.42,2.83 | 0.87 | 0.36,2.11 | 0.86 | 0.36,2.06 | 0.77 | 0.31,1.88 | 0.72 | 0.29,1.76 | 1.4 | 0.48,4.08 | 1.56 | 0.53,4.54 | 1.45 | 0.47,4.42 | 1.38 | 0.44,4.37 |
| **Sex at birth** | | | | | | | | | | | | | | | | | | | | | | | | |
| Female | | | 1.64 | 1.46,1.84 | 1.66 | 1.46,1.88 | 1.79 | 1.58,2.03 | | | 1.57 | 1.39,1.77 | 1.55 | 1.37,1.76 | 1.63 | 1.44,1.86 | | | 1.69 | 1.42,2.01 | 1.65 | 1.39,1.97 | 1.75 | 1.47,2.09 |
| **Ethnicity** | | | | | | | | | | | | | | | | | | | | | | | | |
| White | | | 1 | | 1 | | 1 | | | | 1 | | 1 | | 1 | | | | 1 | | 1 | | 1 | |
| Non-White | | | 1.15 | 0.98,1.34 | 1.11 | 0.93,1.31 | 1.04 | 0.87,1.23 | | | 0.87 | 0.72,1.04 | 0.84 | 0.70,1.00 | 0.79 | 0.66,0.95 | | | 0.88 | 0.69,1.14 | 0.86 | 0.67,1.10 | 0.8 | 0.62,1.04 |
| **Parental income** | | | | | | | | | | | | | | | | | | | | | | | | |
| Lower quintile | | | 1 | | 1 | | 1 | | | | 1 | | 1 | | 1 | | | | 1 | | 1 | | 1 | |
| Q2 | | | 0.95 | 0.78,1.16 | 0.99 | 0.79,1.24 | 1.01 | 0.81,1.28 | | | 0.89 | 0.71,1.12 | 0.91 | 0.72,1.16 | 0.93 | 0.73,1.19 | | | 0.91 | 0.68,1.22 | 0.93 | 0.69,1.26 | 0.96 | 0.71,1.31 |
| Q3 | | | 0.73 | 0.59,0.91 | 0.79 | 0.63,0.99 | 0.86 | 0.68,1.09 | | | 0.83 | 0.66,1.05 | 0.88 | 0.70,1.12 | 0.96 | 0.75,1.23 | | | 0.69 | 0.52,0.91 | 0.72 | 0.55,0.96 | 0.79 | 0.59,1.07 |
| Q4 | | | 0.58 | 0.47,0.70 | 0.61 | 0.49,0.76 | 0.7 | 0.56,0.87 | | | 0.85 | 0.69,1.05 | 0.92 | 0.73,1.14 | 1.04 | 0.83,1.29 | | | 0.49 | 0.38,0.65 | 0.52 | 0.40,0.69 | 0.59 | 0.45,0.79 |
| Highest quintile | | | 0.44 | 0.36,0.53 | 0.47 | 0.38,0.57 | 0.55 | 0.44,0.69 | | | 0.71 | 0.59,0.86 | 0.77 | 0.63,0.95 | 0.9 | 0.73,1.11 | | | 0.43 | 0.32,0.56 | 0.46 | 0.34,0.60 | 0.54 | 0.41,0.72 |
| **Loneliness** | | | | | | | | | | | | | | | | | | | | | | | | |
| Not Lonely | | | | | 1 | | 1 | | | | | | 1 | | 1 | | | | | | 1 | | | 1 |
| Lonely | | | | | 6.08 | 5.36,6.90 | 4.64 | 4.07,5.28 | | | | | 2.92 | 2.58,3.30 | 2.29 | 2.01,2.60 | | | | | 2.45 | 2.06,2.91 | 1.82 | 1.52,2.19 |
| **Support** | | | | | | | | | | | | | | | | | | | | | | | | |
| Yes | | | | | | | 1 | | | | | | | | 1 | | | | | | | | 1 | |
| No | | | | | | | 2.98 | 2.56,3.46 | | | | | | | 2.42 | 2.08,2.82 | | | | | | | 2.62 | 2.15,3.19 |

Model 1 (crude, unadjusted), Model 2: adjusted for ASAB, ethnicity, parental income, Model 3: additionally adjusted for loneliness, and Model 4: additionally adjusted for support. All models included interaction terms between sexual identity and bullying.

**Supplemental Table 6. Predicted probabilities for mental health at age 23 based on past experiences of bullying and sexual identities in 12,782 individuals from the Millennium Cohort Study. Estimates are based on multivariable logistic regression models**

| | Psychological distress | | | | | | Self-harm | | | | | | Attempted suicide | | | | | |
|---|---|---|---|---|---|---|---|---|---|---|---|---|---|---|---|---|---|---|
| Bullying & sexual identity | Model 1 | | | Model 4 | | | Model 1 | | | Model 4 | | | Model 1 | | | Model 4 | | |
| | % | 95% CI | | % | 95% CI | | % | 95% CI | | % | 95% CI | | % | 95% CI | | % | 95% CI | |
| No bullying & Heterosexual | 0.14 | 0.12 | 0.15 | 0.17 | 0.15 | 0.18 | 0.14 | 0.12 | 0.15 | 0.16 | 0.14 | 0.17 | 0.06 | 0.05 | 0.07 | 0.07 | 0.06 | 0.08 |
| No bullying & mainly heterosexual | 0.22 | 0.18 | 0.27 | 0.21 | 0.17 | 0.25 | 0.25 | 0.19 | 0.3 | 0.23 | 0.18 | 0.27 | 0.08 | 0.05 | 0.12 | 0.09 | 0.06 | 0.12 |
| No bullying & Bisexual | 0.36 | 0.28 | 0.43 | 0.29 | 0.23 | 0.35 | 0.43 | 0.36 | 0.51 | 0.37 | 0.31 | 0.44 | 0.19 | 0.13 | 0.25 | 0.17 | 0.12 | 0.23 |
| No bullying & Gay/lesbian | 0.31 | 0.19 | 0.42 | 0.29 | 0.19 | 0.39 | 0.37 | 0.22 | 0.51 | 0.34 | 0.20 | 0.47 | 0.21 | 0.10 | 0.31 | 0.20 | 0.10 | 0.30 |
| No bullying & Other | 0.39 | 0.25 | 0.54 | 0.27 | 0.15 | 0.39 | 0.34 | 0.20 | 0.49 | 0.26 | 0.14 | 0.38 | 0.21 | 0.09 | 0.33 | 0.16 | 0.07 | 0.25 |
| 1 time & Heterosexual | 0.18 | 0.16 | 0.20 | 0.19 | 0.17 | 0.21 | 0.16 | 0.14 | 0.18 | 0.17 | 0.15 | 0.18 | 0.08 | 0.06 | 0.09 | 0.08 | 0.07 | 0.09 |
| 1 time & mainly heterosexual | 0.29 | 0.24 | 0.35 | 0.25 | 0.20 | 0.29 | 0.33 | 0.27 | 0.39 | 0.30 | 0.24 | 0.35 | 0.14 | 0.11 | 0.18 | 0.13 | 0.10 | 0.17 |
| 1 time & Bisexual | 0.39 | 0.31 | 0.47 | 0.32 | 0.25 | 0.38 | 0.47 | 0.39 | 0.54 | 0.40 | 0.33 | 0.47 | 0.33 | 0.26 | 0.40 | 0.29 | 0.22 | 0.36 |
| 1 time & Gay/lesbian | 0.38 | 0.26 | 0.50 | 0.31 | 0.20 | 0.41 | 0.34 | 0.21 | 0.47 | 0.28 | 0.17 | 0.40 | 0.25 | 0.14 | 0.37 | 0.21 | 0.11 | 0.31 |
| 1 time & Other | 0.48 | 0.34 | 0.62 | 0.35 | 0.23 | 0.47 | 0.35 | 0.21 | 0.49 | 0.27 | 0.15 | 0.38 | 0.26 | 0.15 | 0.38 | 0.20 | 0.11 | 0.30 |
| 2 times & Heterosexual | 0.23 | 0.21 | 0.26 | 0.22 | 0.20 | 0.24 | 0.20 | 0.17 | 0.22 | 0.19 | 0.17 | 0.22 | 0.11 | 0.09 | 0.13 | 0.11 | 0.09 | 0.12 |
| 2 times & mainly heterosexual | 0.37 | 0.30 | 0.44 | 0.30 | 0.24 | 0.36 | 0.38 | 0.32 | 0.45 | 0.33 | 0.27 | 0.39 | 0.19 | 0.14 | 0.25 | 0.17 | 0.12 | 0.22 |
| 2 times & Bisexual | 0.45 | 0.36 | 0.54 | 0.31 | 0.24 | 0.38 | 0.49 | 0.39 | 0.58 | 0.38 | 0.30 | 0.46 | 0.29 | 0.21 | 0.36 | 0.22 | 0.16 | 0.28 |
| 2 times & Gay/lesbian | 0.47 | 0.35 | 0.58 | 0.36 | 0.26 | 0.45 | 0.48 | 0.36 | 0.59 | 0.40 | 0.29 | 0.51 | 0.31 | 0.19 | 0.43 | 0.24 | 0.14 | 0.34 |
| 2 times & Other | 0.57 | 0.42 | 0.72 | 0.34 | 0.22 | 0.45 | 0.44 | 0.30 | 0.59 | 0.29 | 0.17 | 0.40 | 0.36 | 0.21 | 0.50 | 0.24 | 0.13 | 0.35 |

Model 1: Unadjusted, Model 4: Adjusted for assigned sex at birth, ethnicity, parental income, loneliness and support. Predicted probabilities based on estimates presented in Supplemental Table Xx.

**Supplemental Table 7. Predicted probabilities for mental health at age 23 based on past experiences of victimisation and sexual identities in 12,782 individuals from the Millennium Cohort Study. Estimates are based on multivariable logistic regression models**

| | Psychological distress | | | | | | Self-harm | | | | | | Attempted suicide | | | | | |
|---|---|---|---|---|---|---|---|---|---|---|---|---|---|---|---|---|---|---|
| **Victimisation & sexual identity** | **Model 1** | | | **Model 4** | | | **Model 1** | | | **Model 4** | | | **Model 1** | | | **Model 4** | | |
| | **%** | **95% CI** | | **%** | **95% CI** | | **%** | **95% CI** | | **%** | **95% CI** | | **%** | **95% CI** | | **%** | **95% CI** | |
| No victimisation & Heterosexual | 0.11 | 0.09 | 0.12 | 0.14 | 0.13 | 0.16 | 0.09 | 0.07 | 0.10 | 0.11 | 0.09 | 0.12 | 0.03 | 0.02 | 0.04 | 0.04 | 0.03 | 0.05 |
| No victimisation & mainly heterosexual | 0.14 | 0.09 | 0.20 | 0.15 | 0.10 | 0.20 | 0.20 | 0.14 | 0.26 | 0.20 | 0.14 | 0.25 | 0.06 | 0.02 | 0.10 | 0.07 | 0.03 | 0.11 |
| No victimisation & Bisexual | 0.25 | 0.15 | 0.35 | 0.21 | 0.13 | 0.28 | 0.29 | 0.20 | 0.39 | 0.26 | 0.17 | 0.35 | 0.07 | 0.01 | 0.13 | 0.06 | 0.01 | 0.11 |
| No victimisation & Gay/lesbian | 0.26 | 0.11 | 0.42 | 0.27 | 0.13 | 0.41 | 0.31 | 0.15 | 0.47 | 0.30 | 0.14 | 0.45 | 0.16 | 0.03 | 0.29 | 0.16 | 0.04 | 0.28 |
| No victimisation & Other | 0.35 | 0.21 | 0.49 | 0.26 | 0.15 | 0.37 | 0.29 | 0.15 | 0.43 | 0.25 | 0.13 | 0.36 | 0.14 | 0.03 | 0.24 | 0.10 | 0.02 | 0.19 |
| 1 time & Heterosexual | 0.15 | 0.14 | 0.17 | 0.17 | 0.15 | 0.19 | 0.15 | 0.13 | 0.16 | 0.16 | 0.14 | 0.18 | 0.06 | 0.05 | 0.08 | 0.07 | 0.06 | 0.08 |
| 1 time & mainly heterosexual | 0.25 | 0.19 | 0.31 | 0.22 | 0.17 | 0.27 | 0.23 | 0.18 | 0.29 | 0.21 | 0.17 | 0.26 | 0.09 | 0.05 | 0.13 | 0.08 | 0.04 | 0.11 |
| 1 time & Bisexual | 0.29 | 0.20 | 0.38 | 0.24 | 0.17 | 0.31 | 0.41 | 0.33 | 0.50 | 0.36 | 0.28 | 0.44 | 0.21 | 0.13 | 0.29 | 0.18 | 0.11 | 0.25 |
| 1 time & Gay/lesbian | 0.29 | 0.14 | 0.43 | 0.25 | 0.13 | 0.37 | 0.24 | 0.10 | 0.39 | 0.22 | 0.10 | 0.35 | 0.17 | 0.06 | 0.29 | 0.15 | 0.06 | 0.25 |
| 1 time & Other | 0.41 | 0.26 | 0.55 | 0.28 | 0.16 | 0.40 | 0.24 | 0.11 | 0.38 | 0.18 | 0.08 | 0.28 | 0.18 | 0.06 | 0.30 | 0.13 | 0.04 | 0.21 |
| ≥2 times & Heterosexual | 0.24 | 0.22 | 0.26 | 0.23 | 0.21 | 0.24 | 0.23 | 0.21 | 0.25 | 0.22 | 0.21 | 0.24 | 0.13 | 0.11 | 0.14 | 0.12 | 0.11 | 0.14 |
| ≥2 times & mainly heterosexual | 0.36 | 0.31 | 0.41 | 0.30 | 0.26 | 0.33 | 0.40 | 0.34 | 0.45 | 0.34 | 0.30 | 0.39 | 0.20 | 0.16 | 0.24 | 0.17 | 0.14 | 0.20 |
| ≥2 times & Bisexual | 0.48 | 0.42 | 0.54 | 0.36 | 0.31 | 0.41 | 0.53 | 0.47 | 0.59 | 0.43 | 0.38 | 0.49 | 0.38 | 0.33 | 0.44 | 0.30 | 0.25 | 0.35 |
| ≥2 times & Gay/lesbian | 0.47 | 0.38 | 0.57 | 0.36 | 0.28 | 0.45 | 0.49 | 0.40 | 0.59 | 0.41 | 0.32 | 0.50 | 0.34 | 0.24 | 0.44 | 0.27 | 0.19 | 0.36 |
| ≥2 times & Other | 0.62 | 0.50 | 0.74 | 0.39 | 0.28 | 0.51 | 0.53 | 0.39 | 0.67 | 0.36 | 0.23 | 0.49 | 0.48 | 0.35 | 0.61 | 0.33 | 0.22 | 0.44 |

Model 1: Unadjusted, Model 4: Adjusted for assigned sex at birth, ethnicity, parental income, loneliness and support. Predicted probabilities based on estimates presented in Supplemental Table Xx.

**Supplemental Table 7. Associations between sexual identity and experiences of cumulative bullying and victimisation across adolescence in complete case sample**

| | Bullying | | | | Victimisation | | | |
|---|---|---|---|---|---|---|---|---|
| | RRR | 95% CI | RRR | 95% CI | RRR | 95% CI | RRR | 95% CI |
| ***No bullying / victimisation*** | | | | | | | | |
| **1 time** | | | | | | | | |
| Heterosexual | 1 | | 1 | | 1 | | 1 | |
| Mainly heterosexual | 1.07 | 0.90,1.28 | 1.11 | 0.93,1.32 | **1.73** | **1.37,2.19** | **1.76** | **1.39,2.23** |
| Bisexual | 1.24 | 0.99,1.56 | 1.26 | 1.00,1.59 | 1.24 | 0.89,1.73 | 1.27 | 0.91,1.78 |
| Gay/lesbian | 1.19 | 0.82,1.74 | 1.20 | 0.82,1.76 | 1.43 | 0.81,2.51 | 1.42 | 0.81,2.50 |
| Other | 1.50 | 0.97,2.33 | 1.49 | 0.95,2.33 | 1.08 | 0.61,1.92 | 1.11 | 0.63,1.96 |
| sex | | | 0.96 | 0.86,1.08 | | | 0.89 | 0.77,1.03 |
| **Ethnicity** | | | | | | | | |
| White | | | 1 | | | | 1 | |
| Non-White | | | 0.92 | 0.78,1.08 | | | **0.77** | **0.64,0.94** |
| **Parental income** | | | | | | | | |
| Lowest quintile | | | 1 | 1 | | | 1 | |
| Q2 | | | 0.95 | 0.77,1.18 | | | **1.42** | **1.10,1.84** |
| Q3 | | | **0.75** | **0.62,0.90** | | | 1 | 0.79,1.27 |
| Q4 | | | **0.73** | **0.61,0.88** | | | 1.17 | 0.93,1.48 |
| Highest quintile | | | **0.68** | **0.57,0.82** | | | 1.05 | 0.83,1.32 |
| **≥2 times** | | | | | | | | |
| Heterosexual | 1 | 1.00,1.00 | 1 | | 1 | | 1 | |
| Mainly heterosexual | **1.42** | **1.13,1.78** | **1.45** | **1.15,1.82** | **2.63** | **2.08,3.32** | **2.72** | **2.15,3.44** |
| Bisexual | **1.54** | **1.21,1.97** | **1.53** | **1.19,1.95** | **2.73** | **2.02,3.69** | **2.84** | **2.09,3.85** |
| Gay/lesbian | **2.57** | **1.70,3.89** | **2.55** | **1.68,3.86** | **2.93** | **1.85,4.65** | **2.92** | **1.85,4.61** |
| Other | **2.58** | **1.61,4.13** | **2.44** | **1.52,3.93** | **1.65** | **1.02,2.66** | **1.72** | **1.06,2.79** |
| sex | | | 1.07 | 0.92,1.23 | | | 0.84 | 0.73,0.97 |
| **Ethnicity** | | | | | | | | |
| White | | | 1 | | | | 1 | |
| Non-White | | | 0.82 | 0.66,1.01 | | | **0.68** | **0.56,0.82** |
| **Parental income** | | | | | | | | |
| Lowest quintile | | | 1 | | | | 1 | |
| Q2 | | | **1.39** | **1.08,1.78** | | | **1.53** | **1.16,2.02** |

| | | | | | | | | |
|---|---|---|---|---|---|---|---|---|
| Q3 | | | 0.95 | 0.76,1.18 | | | 1.22 | 0.93,1.62 |
| Q4 | | | 0.85 | 0.67,1.08 | | | 1.18 | 0.91,1.53 |
| Highest quintile | | | **0.69** | **0.54,0.87** | | | 1.04 | 0.80,1.36 |
| ***Observations*** | ***8596*** | | ***8586*** | | ***6975*** | | ***6968*** | |

**Supplemental Table 8. Associations between frequency of bullying in adolescence and mental health outcomes in early adulthood in complete case sample**

| | Bullying | | | | | | | Victimisation | | | | | |
|---|---|---|---|---|---|---|---|---|---|---|---|---|---|
| | **Psychological distress** | | **Self-harm** | | **Attempted suicide** | | | **Psychological distress** | | **Self-harm** | | **Attempted suicide** | |
| | **OR** | **95% CI** | **OR** | **95% CI** | **OR** | **95% CI** | | **OR** | **95% CI** | **OR** | **95% CI** | **OR** | **95% CI** |
| **Bullying** | | | | | | | **Victimisation** | | | | | | |
| None | 1 | | 1 | | 1 | | None | 1 | | 1 | | 1 | |
| 1 time | 1.21 | 0.96,1.53 | 1.03 | 0.84,1.26 | 1.02 | 0.77,1.35 | 1 time | 1.24 | 0.95,1.62 | **1.83** | **1.43,2.35** | **2.32** | **1.54,3.48** |
| 2 times | **1.57** | **1.24,1.97** | **1.31** | **1.05,1.64** | **1.46** | **1.07,1.99** | 2/3 times | **1.87** | **1.52,2.29** | **2.83** | **2.24,3.58** | **3.85** | **2.68,5.51** |
| **Sexual identity** | | | | | | | **Sexual identity** | | | | | | |
| Heterosexual | 1 | | 1 | | 1 | | Heterosexual | 1 | | 1 | | 1 | |
| Mainly heterosexual | 1.33 | 0.96,1.84 | 1.45 | 1.06,1.99 | 0.89 | 0.55,1.44 | Mainly heterosexual | 0.86 | 0.51,1.46 | 2.61 | 1.52,4.47 | 1.81 | 0.74,4.44 |
| Bisexual | **2.59** | **1.65,4.06** | **3.80** | **2.61,5.55** | **2.50** | **1.51,4.12** | Bisexual | **1.73** | **0.92,3.25** | **3.69** | **2.04,6.70** | **1.15** | **0.40,3.28** |
| Gay/lesbian | **2.98** | **1.46,6.08** | **3.77** | **1.81,7.84** | **3.90** | **1.66,9.16** | Gay/lesbian | **3.84** | **1.40,10.53** | **6.16** | **2.34,16.21** | **7.68** | **2.59,22.83** |
| Other | 2.68 | 0.92,7.82 | 2.40 | 0.99,5.85 | 1.77 | 0.69,4.54 | Other | **4.44** | **1.88,10.47** | **5.50** | **2.52,12.00** | 3.23 | 0.92,11.39 |
| **Interaction terms for bullying X sexual identity** | | | | | | | **Interaction terms for victimisation X sexual identity** | | | | | | |
| No bullying | 1 | | 1 | | 1 | | No victimisation | 1 | | 1 | | 1 | |
| bully_total=1 ~o | 1.24 | 0.76,2.02 | **1.77** | **1.09,2.86** | **2.15** | **1.13,4.10** | bully_total=1 ~o | **1.85** | **1.02,3.33** | **0.52** | **0.27,0.98** | 0.46 | 0.15,1.42 |
| bully_total=1 ~l | 0.90 | 0.46,1.76 | 1.09 | 0.63,1.89 | **2.43** | **1.24,4.75** | bully_total=1 ~l | 0.92 | 0.38,2.22 | 1.03 | 0.47,2.26 | 2.74 | 0.79,9.48 |
| bully_total=1 ~b | 0.74 | 0.24,2.30 | 0.37 | 0.11,1.21 | 0.71 | 0.19,2.63 | bully_total=1 ~b | 0.56 | 0.15,2.17 | **0.21** | **0.06,0.75** | 0.31 | 0.08,1.16 |
| bully_total=1 ~r | 1.61 | 0.44,5.92 | 0.81 | 0.26,2.51 | 1.88 | 0.53,6.60 | bully_total=1 ~r | 0.57 | 0.16,2.04 | **0.15** | **0.05,0.47** | 0.22 | 0.03,1.49 |
| bully_total=2 ~o | 1 | | 1 | | 1 | | bully_total=2 ~o | 1 | | 1 | | 1 | |
| bully_total=2 ~o | 1.44 | 0.89,2.33 | 1.51 | 0.94,2.41 | **2.11** | **1.04,4.28** | bully_total=2 ~o | 1.88 | 0.99,3.54 | 0.75 | 0.40,1.39 | 0.87 | 0.34,2.25 |
| bully_total=2 ~l | 0.69 | 0.35,1.39 | 0.68 | 0.36,1.30 | 0.80 | 0.39,1.65 | bully_total=2 ~l | 1.38 | 0.69,2.76 | 0.78 | 0.39,1.56 | 2.96 | 1.00,8.72 |
| bully_total=2 ~b | 0.88 | 0.35,2.19 | 0.73 | 0.28,1.87 | 0.62 | 0.20,1.92 | bully_total=2 ~b | 0.64 | 0.19,2.14 | 0.41 | 0.14,1.22 | 0.35 | 0.11,1.15 |
| bully_total=2 ~r | 0.62 | 0.17,2.30 | 0.50 | 0.16,1.50 | 0.95 | 0.26,3.50 | bully_total=2 ~r | 0.61 | 0.19,1.95 | **0.34** | **0.12,0.94** | 1.15 | 0.28,4.76 |
| **Sex** | | | | | | | **Sex** | | | | | | |
| Male | 1 | | 1 | | 1 | | Male | 1 | | 1 | | 1 | |
| Female | **1.67** | **1.43,1.95** | **1.66** | **1.42,1.93** | **1.53** | **1.26,1.85** | Female | **1.74** | **1.49,2.02** | **1.66** | **1.43,1.92** | **1.54** | **1.25,1.89** |
| **Ethnicity** | | | | | | | **Ethnicity** | | | | | | |

| | | | | | | | | | | | | | |
|---|---|---|---|---|---|---|---|---|---|---|---|---|---|
| White | 1 | | 1 | | 1 | | White | 1 | | 1 | | 1 | |
| Non-White | 0.99 | 0.80,1.23 | **0.74** | **0.60,0.90** | 0.77 | 0.55,1.08 | Non-White | 1.04 | 0.84,1.29 | **0.79** | **0.65,0.96** | 0.90 | 0.66,1.22 |
| **Parental income** | | | | | | | **Parental income** | | | | | | |
| Lowest quintile | 1 | | 1 | | 1 | | Lowest quintile | 1 | | 1 | | 1 | |
| Q2 | 0.92 | 0.69,1.21 | 0.83 | 0.62,1.11 | 1.08 | 0.75,1.56 | Q2 | 0.94 | 0.71,1.24 | 0.79 | 0.60,1.04 | 0.95 | 0.67,1.36 |
| Q3 | 0.91 | 0.71,1.18 | 0.86 | 0.64,1.17 | 1.05 | 0.76,1.45 | Q3 | 0.83 | 0.65,1.07 | 0.88 | 0.66,1.18 | 0.93 | 0.68,1.27 |
| Q4 | **0.71** | **0.55,0.92** | 1.03 | 0.79,1.33 | **0.66** | **0.48,0.92** | Q4 | **0.66** | **0.51,0.85** | 0.96 | 0.75,1.24 | **0.57** | **0.42,0.79** |
| Highest quintile | **0.54** | **0.41,0.70** | 0.85 | 0.65,1.12 | **0.64** | **0.46,0.89** | Highest quintile | **0.51** | **0.39,0.65** | 0.80 | 0.62,1.05 | **0.56** | **0.41,0.77** |
| **Loneliness** | | | | | | | **Loneliness** | | | | | | |
| Not lonely | 1 | | 1 | | 1 | | Not lonely | 1 | | 1 | | 1 | |
| Lonely | **4.30** | **3.65,5.07** | **2.32** | **1.97,2.73** | **2.01** | **1.62,2.50** | Lonely | **4.22** | **3.60,4.96** | **2.18** | **1.86,2.55** | **1.85** | **1.50,2.28** |
| **Support** | | | | | | | **Support** | | | | | | |
| Yes | 1 | | 1 | | 1 | | Yes | 1 | | 1 | | 1 | |
| No | **3.21** | **2.67,3.85** | **2.6** | **2.20,3.08** | **2.69** | **2.13,3.40** | No | **3.14** | **2.64,3.74** | **2.52** | **2.13,2.97** | **2.52** | **2.01,3.16** |
| *Observations* | *6648* | | *6653* | | *6643* | | *Observations* | *6933* | | *6938* | | *6928* | |
